# What is the evidence for transmission of COVID-19 by children in schools? A living systematic review

**DOI:** 10.1101/2020.10.11.20210658

**Authors:** Wei Xu, Xue Li, Marshall Dozier, Yazhou He, Amir Kirolos, Zhongyu Lang, Catherine Mathews, Nandi Siegfried, Evropi Theodoratou, on behalf of UNCOVER

## Abstract

**Background:** It is of paramount importance to understand the transmission of SARS-CoV-2 in schools, which could support the decision-making about educational facilities closure or re-opening with effective prevention and control measures in place.

**Methods:** We conducted a systematic review and meta-analysis to investigate the extent of SARS-CoV-2 transmission in schools. We performed risk of bias evaluation of all included studies using the Newcastle-Ottawa Scale (NOS).

**Results:** 2,178 articles were retrieved and 11 studies were included. Five cohort studies reported a combined 22 student and 21 staff index cases that exposed 3,345 contacts with 18 transmissions [overall infection attack rate (IAR): 0.08% (95% CI: 0.00%–0.86%)]. IARs for students and school staff were 0.15% (95% CI: 0.00%–0.93%) and 0.70% (95% CI: 0.00%–3.56%) respectively. Six cross-sectional studies reported 639 SARS-CoV-2 positive cases in 6,682 study participants tested [overall SARS-CoV-2 positivity rate: 8.00% (95% CI: 2.17%–16.95%)]. SARS-CoV-2 positivity rate was estimated to be 8.74% (95% CI: 2.34%–18.53%) among students, compared to 13.68% (95% CI: 1.68%–33.89%) among school staff. Gender differences were not found for secondary infection (OR: 1.44, 95% CI: 0.50-4.14, P= 0.49) and SARS-CoV-2 positivity (OR: 0.90, 95% CI: 0.72-1.13, P= 0.36) in schools. Fever, cough, dyspnea, ageusia, anosmia, rhinitis, sore throat, headache, myalgia, asthenia, and diarrhoea were all associated with the detection of SARS-CoV-2 antibodies (based on two studies). Overall, study quality was judged to be poor with risk of performance and attrition bias, limiting the confidence in the results.

**Conclusions:** There is limited high-quality evidence available to quantify the extent of SARS-CoV-2 transmission in schools or to compare it to community transmission. Emerging evidence suggests lower IAR and SARS-CoV-2 positivity rate in students compared to school staff. Future prospective and adequately controlled cohort studies are necessary to confirm this finding.

## INTRODUCTION

Globally, there have been at least 29,737,453 confirmed Coronavirus Disease 2019 (COVID-19) cases and 937,391 deaths have occurred in 216 countries/territories according to the report of WHO from 17 September 2020 [1]. In response to the pandemic of novel COVID-19 caused by a severe acute respiratory syndrome coronavirus 2 (SARS-CoV-2), 107 countries had implemented national school closures by March 18 2020 to reduce transmission [2].

Initial evidence suggests children have lower susceptibility and relatively small proportion of infections, compared to adults [3]. Children also have milder cases and better prognosis than adults [4]. According to data from 29 countries, the proportion of children among COVID-19 cases varies from 0.3% (lowest in Spain) up to 13.8% (highest in Argentina) [5].

Many schools closed at the beginning of the COVID-19 pandemic and therefore it is not known whether children are at risk of higher transmission in school settings compared to community settings. Multiple countries around the world have now re-opened schools for face-to-face teaching with varying non-pharmaceutical interventions (NPIs) in place including physical distancing measures, wearing of face masks, enhanced hand hygiene, reduced class sizes, and staggered class start and end times [6]. Evidence on SARS-CoV-2 transmission in schools could support decision-making about schools/childcare facilities closure or re-opening with effective COVID-19 prevention and control measures in place.

A living systematic review to investigate the evidence of SARS-CoV-2 transmission in the school environment is presented. We aim to keep updating this systematic review to include new studies as they become available and to re-evaluate the conclusions given the rapid pace of ongoing research.

## METHODS

### Protocol

The protocol of this living systematic review was developed in accordance with the reporting guidance in the Preferred Reporting Items for Systematic Reviews and Meta-Analyses Protocols (PRISMA-P) statement [7] and was registered on PROSPERO (register number: CRD42020192839) [8].

### Literature search and eligibility criteria

We ran a systematic search in MEDLINE, CINAHL, ERIC, Embase, WHO COVID-19 database, medRxiv, The American Academy of Pediatrics (AAP), The Royal College of Paediatrics and Child Health (RCPCH), and *Do not forget the bubbles* websites with entry date limits from December 2019 to 14 July 2020 (please see search strategies in Appendix S1 of the Online Supplementary Document), to identify studies that investigated SARS-CoV-2 transmission in schools. We ran an updated search in MEDLINE up to 14 September 2020. We further hand-searched reference lists of the retrieved eligible publications to identify additional relevant studies. We reviewed titles, abstracts, and subsequently full texts based on pre-defined inclusion and exclusion criteria following the population, exposure, comparison, outcome (PECO) approach. We included children (defined as ≤18 years old) who were attending school, and their close contacts (family and household members, teachers, school support staff) during the COVID-19 pandemic. We excluded home-schooled children and their close contacts and schools with student numbers below 20. For study outcomes, we included infections traced to a school index case with a COVID-19 positive test. For study types, inclusion criteria spanned cohort studies regardless of active or passive follow-up in the exposed and non-exposed groups (e.g. contact-tracing studies), viral genotyping studies, cross-sectional studies (e.g. sero-surveillance studies, community prevalence studies before and after school opening). We included articles in peer-reviewed journals and pre-prints, and excluded comments, conference abstracts and interviews.

### Data extraction

Data relevant to the evidence for SARS-CoV-2 transmission in schools were extracted including: citation details, publication type, study design, country, region, city, investigation period, background population setting (country/regional COVID-19 prevalence rates where reported), types of non-pharmaceutical intervention in the background population setting, school closures at the time of the study, number of schools included, type of schools, size of schools, types of non-pharmaceutical interventions in place in schools, sampling method (nasopharyngeal or oropharyngeal swabs/ serum samples), provider testing versus self-testing, testing method (PCR/ SARS-CoV-2 antibody testing), modality of follow-up, frequency of follow-up, case and contact demographics (age and gender), clinical characteristics, number of index cases, number of contacts, number of secondary infected cases, infection attack rates (IAR): no. of secondary infected cases/ No. of contacts, number of participants tested for SARS-CoV-2, number of SARS-CoV-2 positive cases, and SARS-CoV-2 positivity rates: no. of positive cases/ No. of participants tested. Data were extracted by one reviewer (WX) and checked by a second reviewer (YH).

### Meta-analysis

We pooled together SARS-CoV-2 infection attack rates (IAR) and positivity rates using a random-effects model (DerSimonian-Laird) [9]. To account for zero cell counts, we transformed raw numbers/proportions with the Freeman-Tukey double arcine method to stabilize the variance [10].

We performed further random-effects meta-analyses (DerSimonian-Laird) of the association of SARS-CoV-2 positivity with gender and clinical symptoms. Symptoms were further categorized as major (fever, cough, dyspnoea, anosmia and ageusia) or minor (sore throat, rhinitis, myalgia, diarrhoea, headache, asthenia) [22-23].

Heterogeneity among studies was tested using Cochran’s Q statistic, the I^2^ index, and the tau-squared test [11]. Funnel plots and the Egger test were used to detect evidence of publication bias [12]. P < 0.05 was considered as statistically significant (two-sided).

### Assessment of methodological quality and risk of bias

We applied the Newcastle Ottawa Scale (NOS) for controlled cohort studies to reflect the school setting [43] and used the NOS as a foundation to evaluate the quality of cross-sectional studies informed by earlier work [44]. The tools included an assessment of selection, measurement and attrition bias, and comparability. The tool is available in the supplementary materials (Appendix S2 of the Online Supplementary Document).

All statistical analyses were conducted using R, version 3.3.0 (R Foundation for Statistical Computing).

## RESULTS

The initial search retrieved 2,178 articles. After screening, 11 studies were eligible for inclusion (Figure 1), including five cohort studies [13-17] and six cross-sectional studies [18-23]. We did not identify viral genotyping studies.

**Figure 1:**
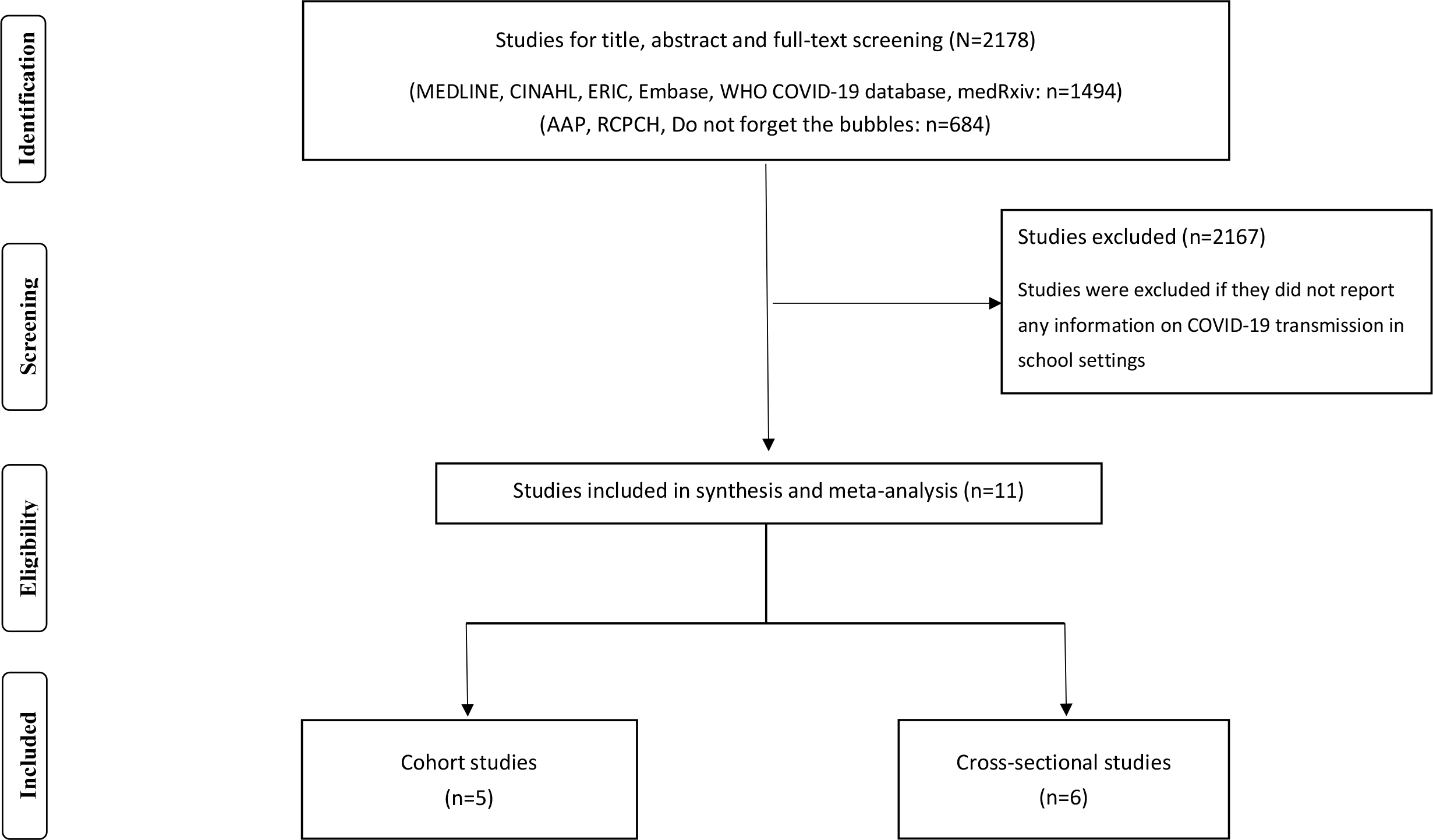
Flowchart summarizing study identification and selection

### Characteristics and quality of the included studies

The study characteristics of the 11 included studies are presented in **Tables 1 and 2**.

**Table 1:**
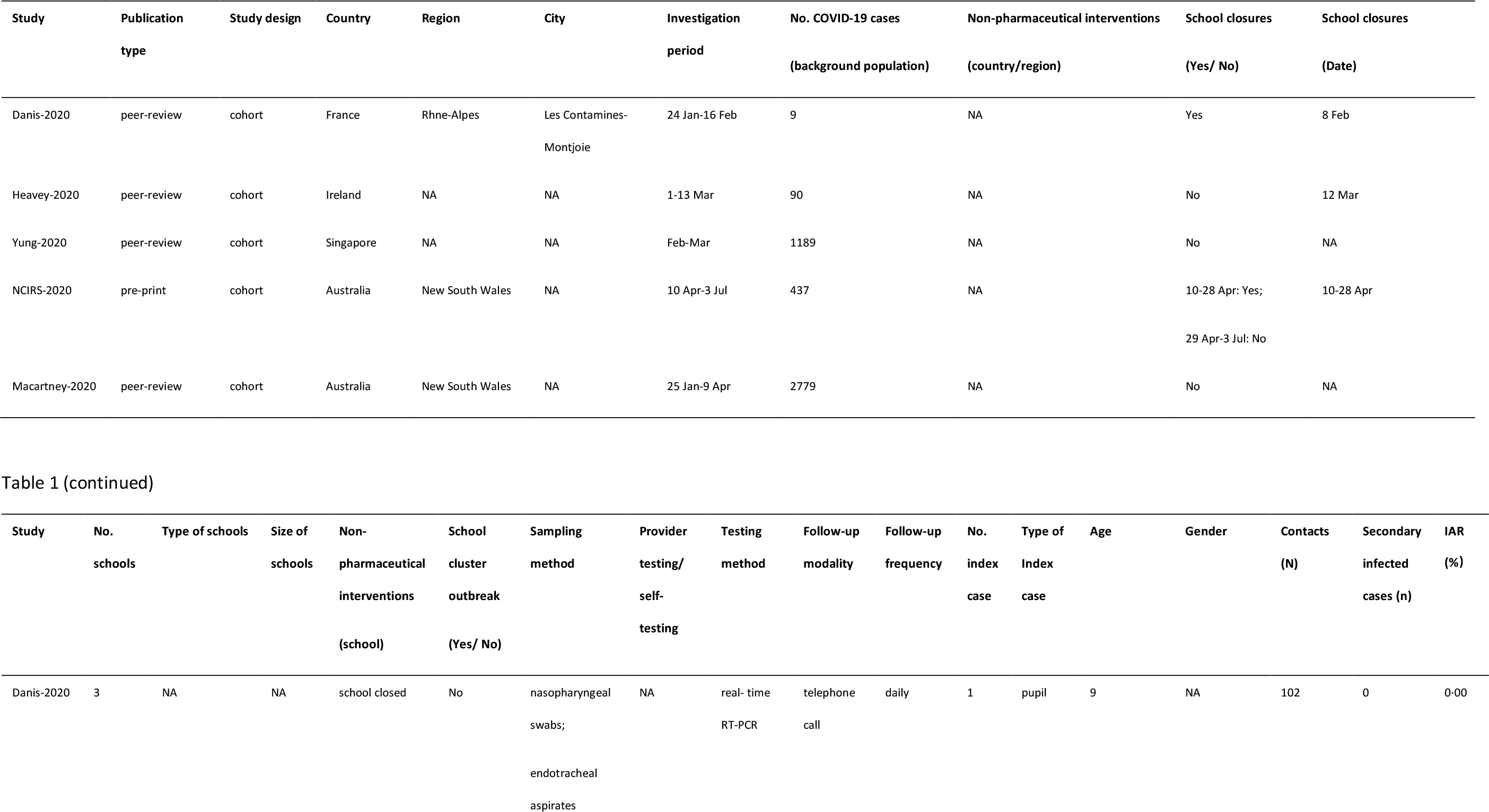
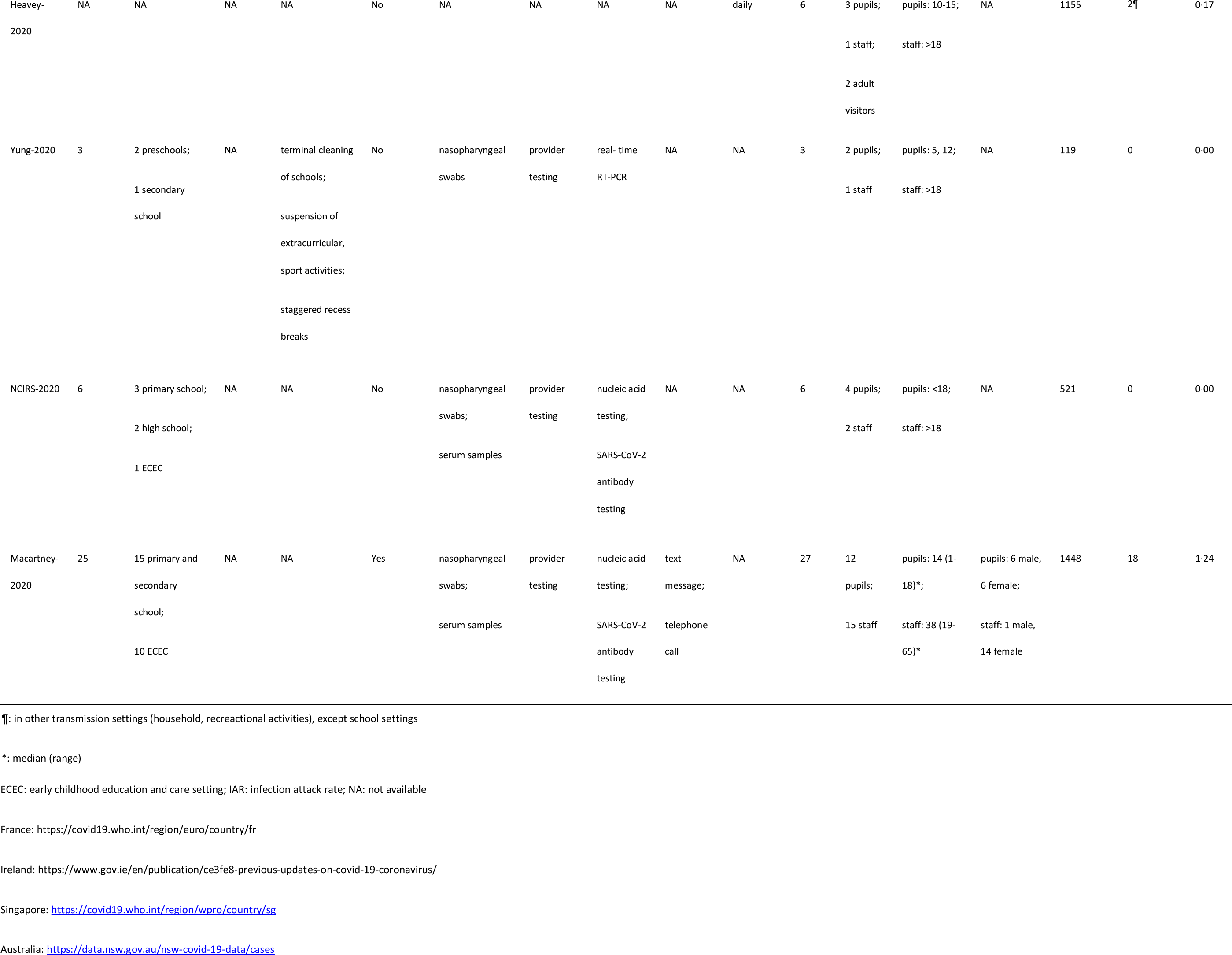
Characteristics of cohort studies (N=5)

**Table 2:**
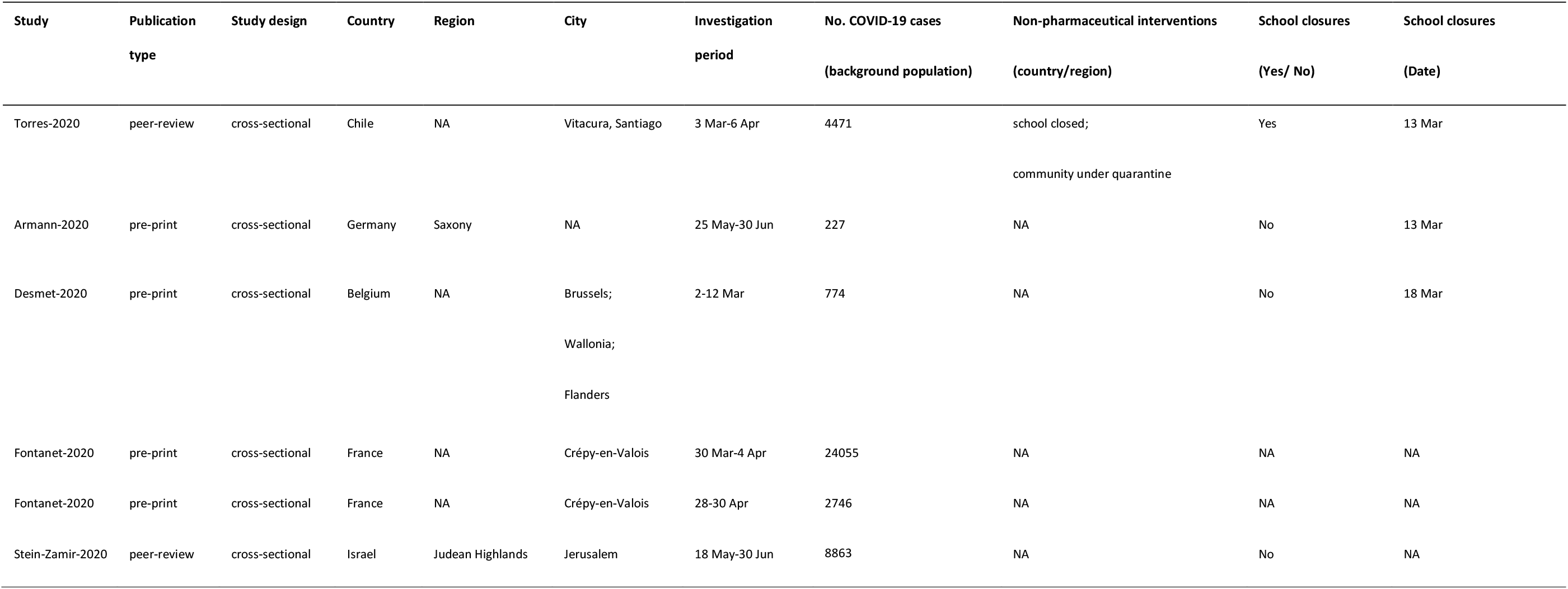
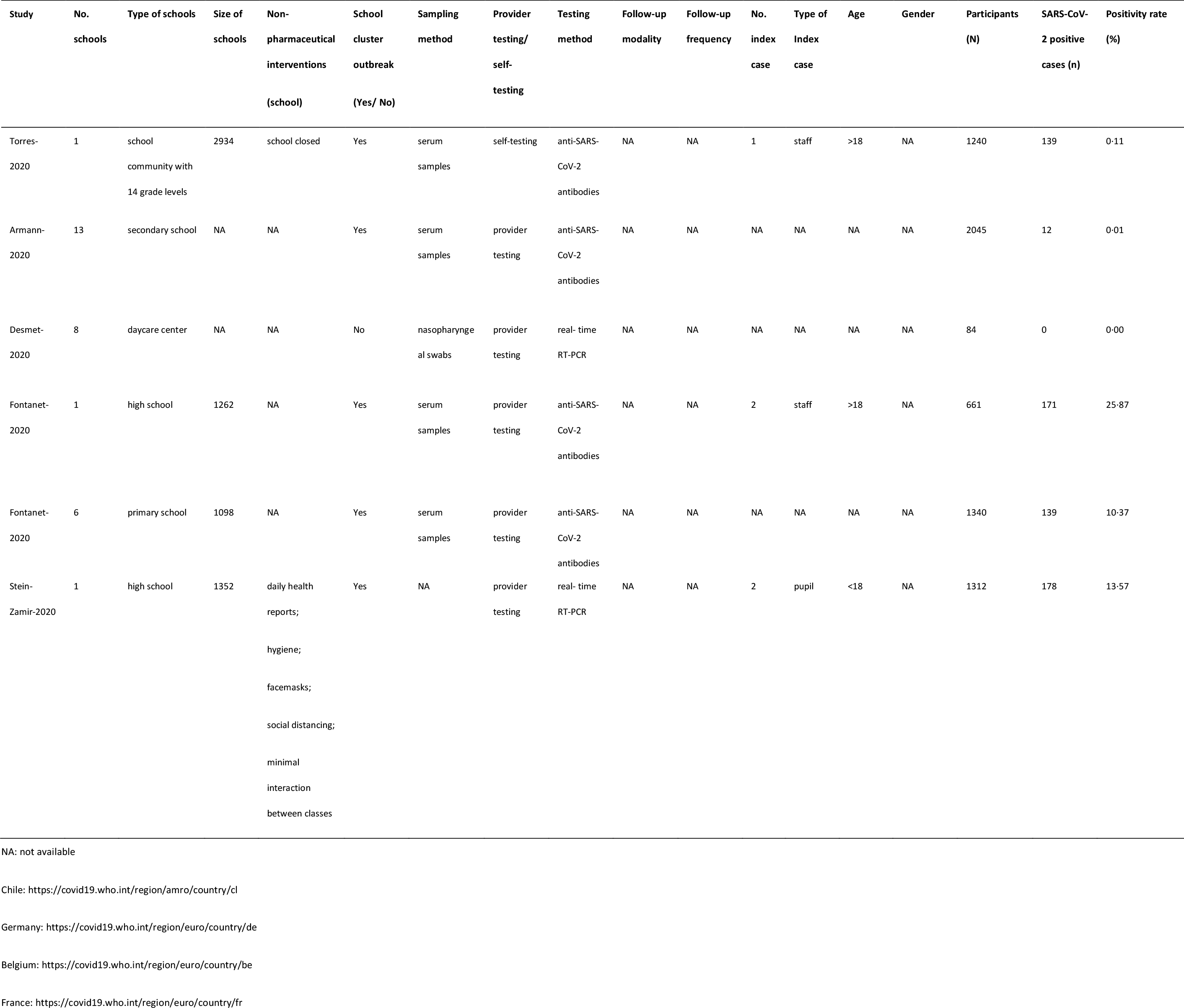
Characteristics of cross-sectional studies (N=6)

#### Cohort studies

A cluster outbreak in schools was reported in Australia New South Wales (NSW) during 25 January-9 April [17]. In NSW, 15 primary and secondary schools, and ten early childhood education and care (ECEC) settings had 27 primary SARS-CoV-2 positive cases including 12 children and 15 school staff attending while infectious, with 1,448 contacts traced [17]. Secondary transmission was reported in three schools and one ECEC. Eighteen secondary infected cases were found among a total of 1,448 close contacts. IARs for primary school, secondary school, ECDC and overall were 0.92%, 0.00%, 2.25% and 1.24% respectively. Transmission rate of student-to-student was 0.31%, and student-to-school staff was 0.97%. By comparison, transmission rate of school staff-to-student was 1.49% and school staff-to-school staff was 4.38%.

The remaining three studies in France (Les Contamines-Montjoie), Ireland, and Singapore and a follow-up of the NSW Australian study indicated that the extent of any student-to-student and/or student-to-school staff transmission is limited [13-16].

In France (Les Contamines-Montjoie), a 9-year-old child attended three different schools while symptomatic, and of the 102 contacts identified, no secondary infections occurred [13].

A study in Ireland investigated SARS-CoV-2 transmission in schools before school closures on 12 March and did not identify any cases of onward transmission to other students or school staff [14]. In this study, 6 primary COVID-19 cases including three students, one teacher and two adult visitors who attended educational sessions were identified and 1,155 contacts (924 student contacts and 101 adult contacts) were identified.

During February and March, nationwide surveillance and contact-tracing in Singapore identified two SARS-CoV-2 positive students (5-year-old and 12-year-old) who attended pre-school and secondary school on the first day of their symptoms before subsequently being diagnosed with COVID-19, and one school staff who worked in a pre-school [15]. Screening of 119 students and staff who were close contacts (secondary school: n=8; pre-school 1: n=34; pre-school 2: n=77) did not detect any SARS-CoV-2 infection.

In the NSW follow-up study, (school term 2 of the academic year between 10 April and 3 July), six SARS-CoV-2 positive cases including four students and two school staff attended three primary schools, two high schools and one ECEC while infectious, and 521 contacts (459 student contacts and 62 adult contacts) were identified [16]. No secondary infection was reported.

#### Cross-sectional studies

A study in Belgium measured the prevalence of SARS-CoV-2 in randomly sampled 84 children attending eight daycare centres during the period 2-12 March, and found all analyzed samples were negative for SARS-CoV-2 [21].

Four studies in Chile (Vitacura, Santiago), Germany (Saxony), and France (Crépy-en-Valois) identified antibody positive cases in schools, and overall seroprevalence varied from 0.01% to 25.87% [19-20, 22-23]. A large school community was closed on 13 March in Chile (Santiago) and during quarantine, a home-delivery and self-administered antibody test were conducted among 1,009 students and 235 school staff [19]. Antibody positive rates were 9.91% (100/1009) and 16.60% (39/235) respectively. Antibody positive rates for pre-school, elementary school, middle school and high school were 12.24%, 10.84%, 11.80% and 5.69%. The peak rate was observed in pre-school.

After reopening of schools in Germany (Saxony) on 18 May, 1,538 students from grade 8–11 and 507 teachers in 13 secondary schools were tested for SARS-CoV-2 antibody to investigate their role in SARS-CoV-2 transmission in schools [20]. The overall antibody positive rate was 0.58% (12/2045), and 0.72% (11/1538) for student and 0.20% (1/507) for school staff.

In France (Crépy-en-Valois), two sero-prevalence studies were conducted between 30 March-30 April in one high school (n=661) and six primary schools (n=1340) [22-23]. Antibody positive rates were 25.87% (171/661) in the high school and 10.37% (139/1340) in primary schools. Specifically, seropositivity prevalence was 38.33% (92/240), 48.75% (39/80) among students and staff in high school. By comparison, seropositivity prevalence was 8.82% (45/510), 5.71% (4/70) among students and staff in primary schools.

In Israel, ten days after schools reopened on 17 May, two index student cases were reported in a high school [18]. SARS-CoV-2 real-time PCR tests were provided to 1,161 students and 151 school staff, a total of 178 positive cases (overall positivity: 13.57%) including 153 students (student positivity: 13.18%) and 25 school staff (staff positivity: 16.56%) were identified. SARS-CoV-2 positive rates were higher in junior grades for students aged 12-14 years old than in high grades for students aged 15-18 years old. The peak rates were observed in the 9th grade (14 year old, 32.62%) and the 7th grade (12 year old, 20.30%).

### SARS-CoV-2 infection attack rate

We combined SARS-CoV-2 IARs in schools in meta-analyses (Table 3). A total of five cohort studies early in pandemic before lockdown were included with 18 secondary infected cases in 3,345 contacts [13-17]. The pooled IAR of total study participants was calculated to be 0.08% (95% CI: 0.00%–0.86%) by using the Freeman-Tukey double arcine transformation and DerSimonian-Laird random-effects model (Figure 2A). The heterogeneity in this meta-analysis was substantial with an I^2^ value of 86.2%. There was no evidence of publication bias (Egger’s test p = 0.661; Figure 2B, 2C).

**Table 3:** SARS-CoV-2 infection attack rate meta-analyses results.

|  | Number of<br>studies | n (infected cases) | N (contacts) | IAR (%) | 95% CI | Cochrane Q | I<sup>2</sup> | Tau-square | P-Egger |
| --- | --- | --- | --- | --- | --- | --- | --- | --- | --- |
| Total | 5 | 18 | 3345 | 0.08 | 0.00-0.86 | 29.06 | 86.20 | 0.0028 | 0.6611 |
| Student | 3 | 10 | 2568 | 0.15 | 0.00-0.93 | 14.61 | 86.30 | 0.0020 | 0.5801 |
| School staff | 3 | 8 | 426 | 0.70 | 0.00-3.56 | 6.40 | 68.80 | 0.0047 | 0.2572 |

CI: confidence interval; IAR: infection attack rate

**Figure 2A:**
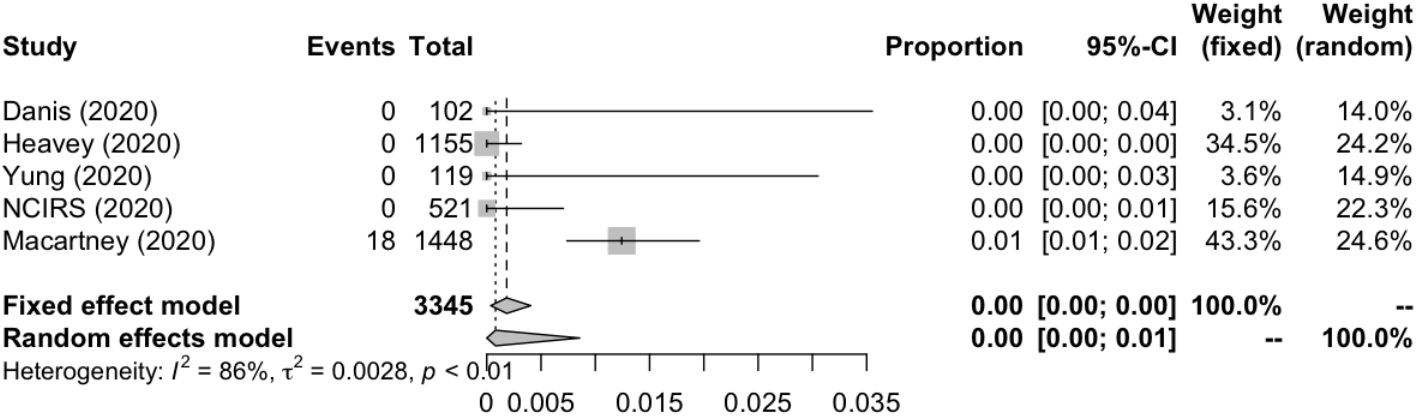
Forest plot (overall infection attack rate)

**Figure 2B:**
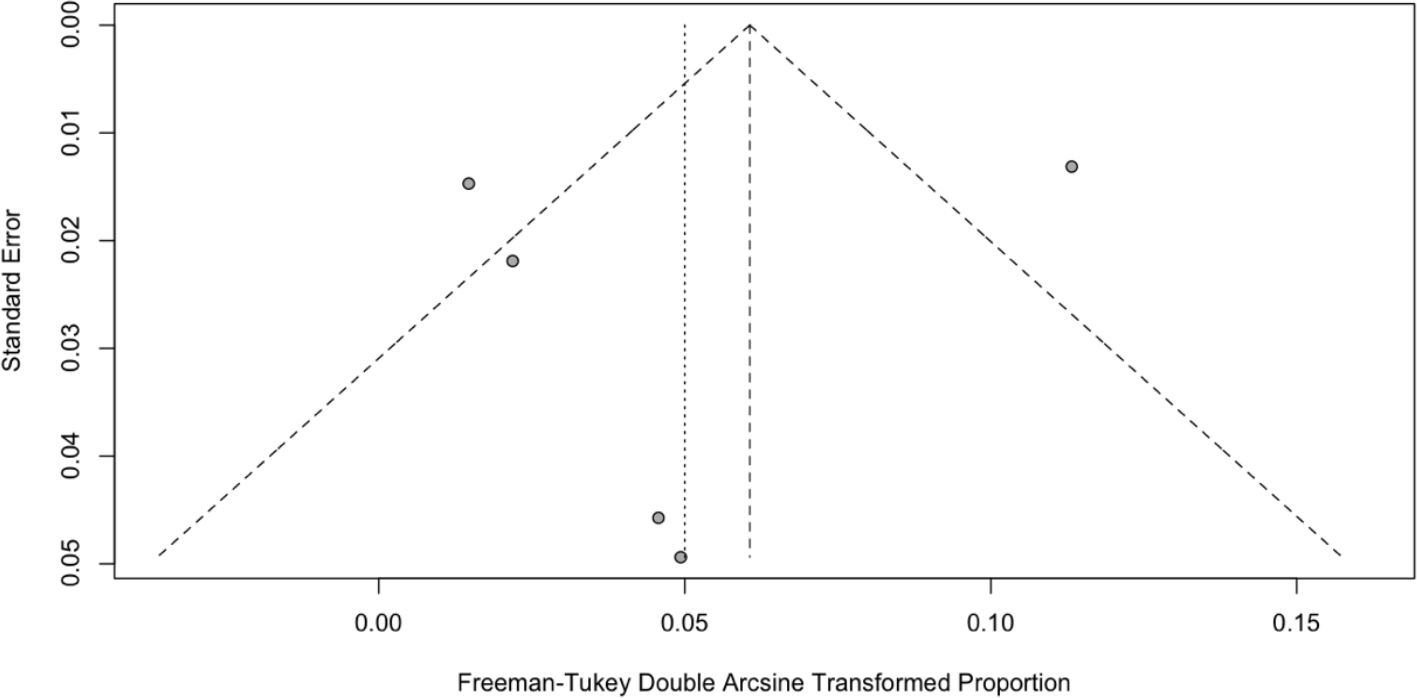
Funnel plot (overall infection attack rate)

**Figure 2C:**
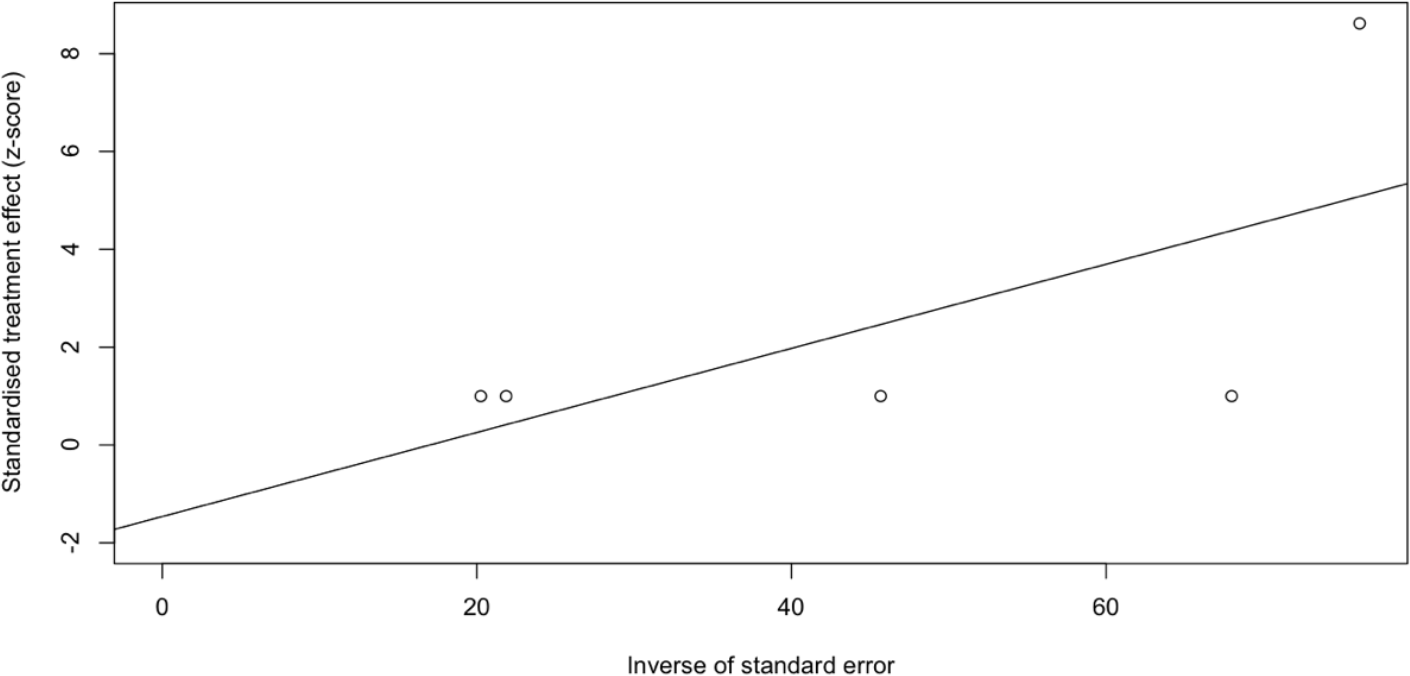
Egger’s publication bias plot (overall infection attack rate)

We estimated the pooled IARs for students and school staff separately, [0.15% (95% CI: 0.00%–0.93%) and 0.70% (95% CI: 0.00%–3.56%) respectively; Figure 3A and 4A]. Heterogeneity was high and there was no evidence of publication bias (Figure 3B, 3C and 4B, 4C).

**Figure 3A:**
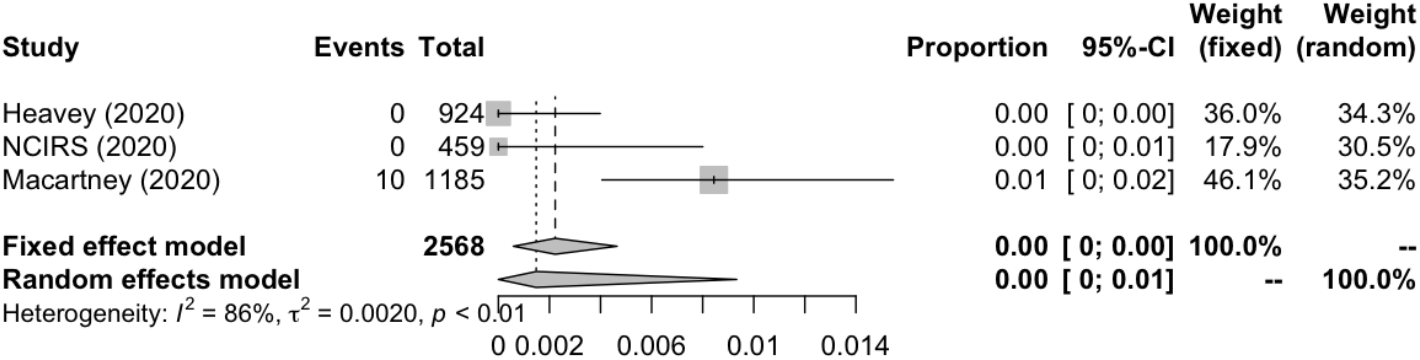
Forest plot (student infection attack rate)

**Figure 3B:**
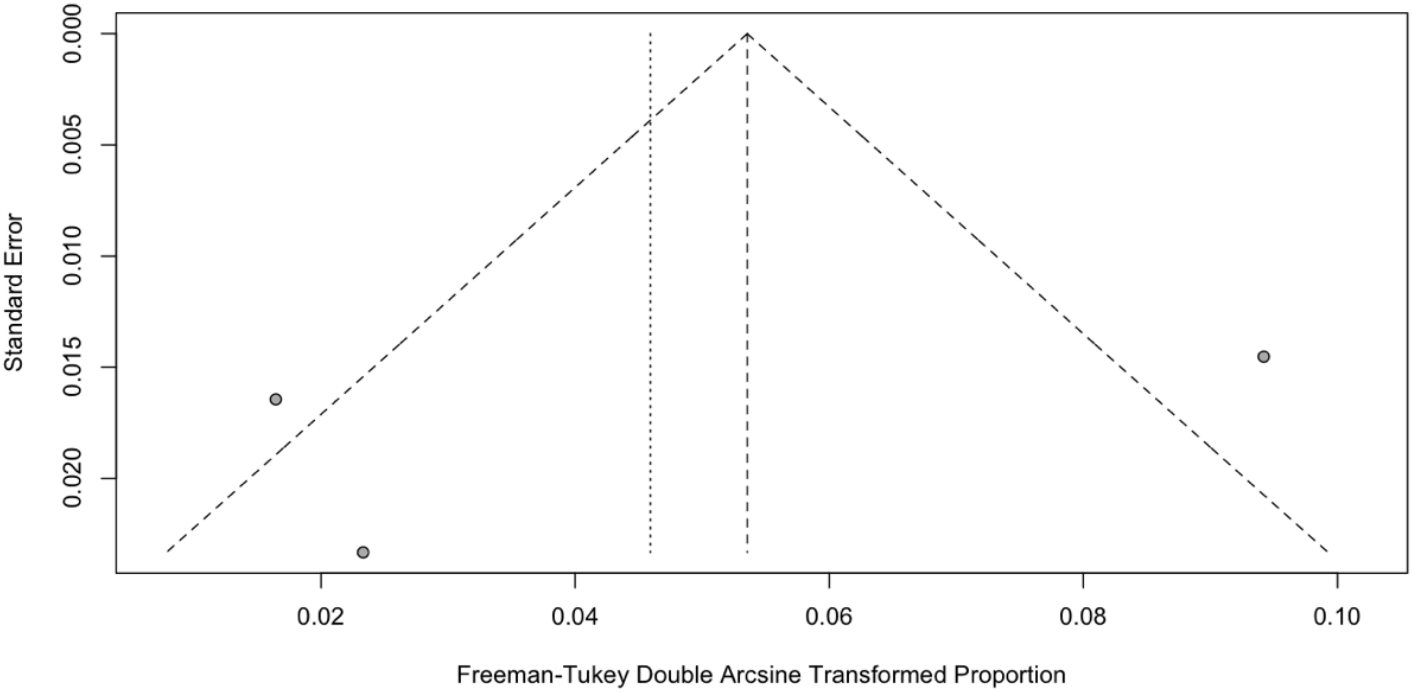
Funnel plot (student infection attack rate)

**Figure 3C:**
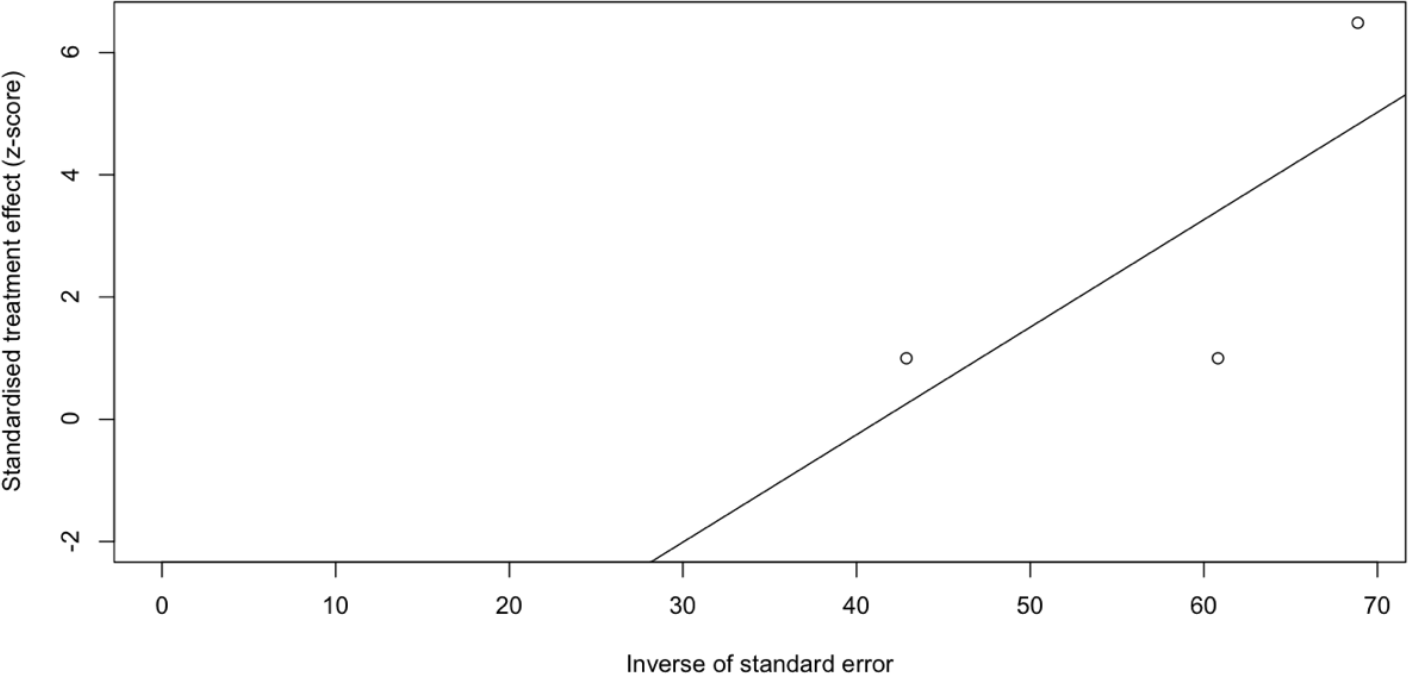
Egger’s publication bias plot (student infection attack rate)

**Figure 4A:**
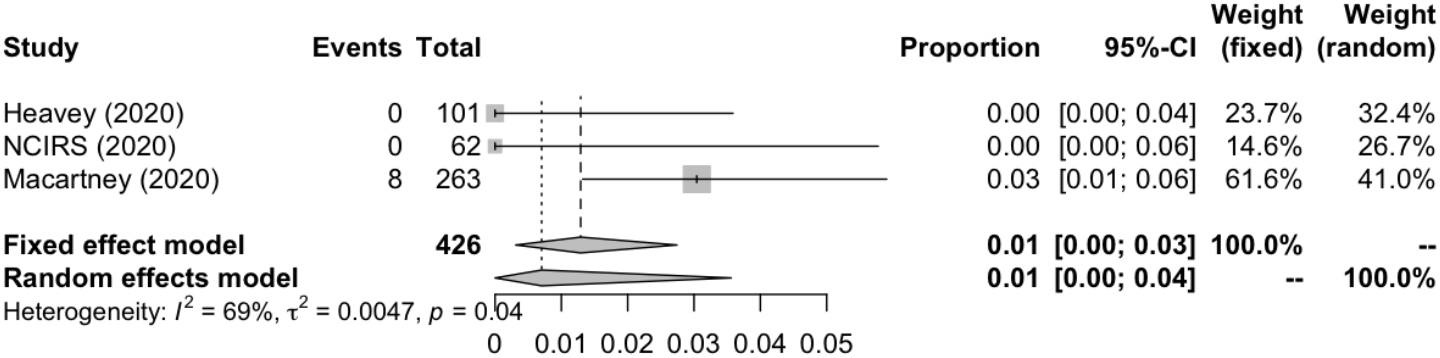
Forest plot (school staff infection attack rate)

**Figure 4B:**
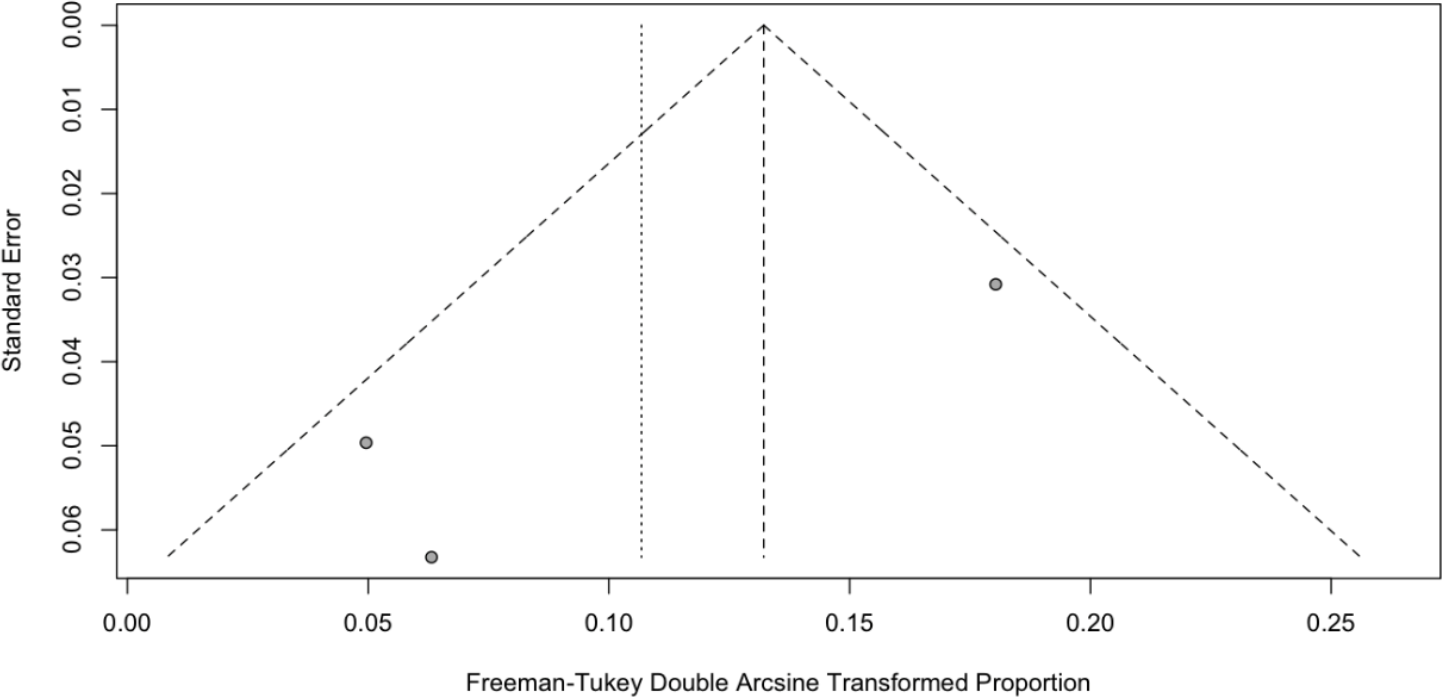
Funnel plot (school staff infection attack rate)

**Figure 4C:**
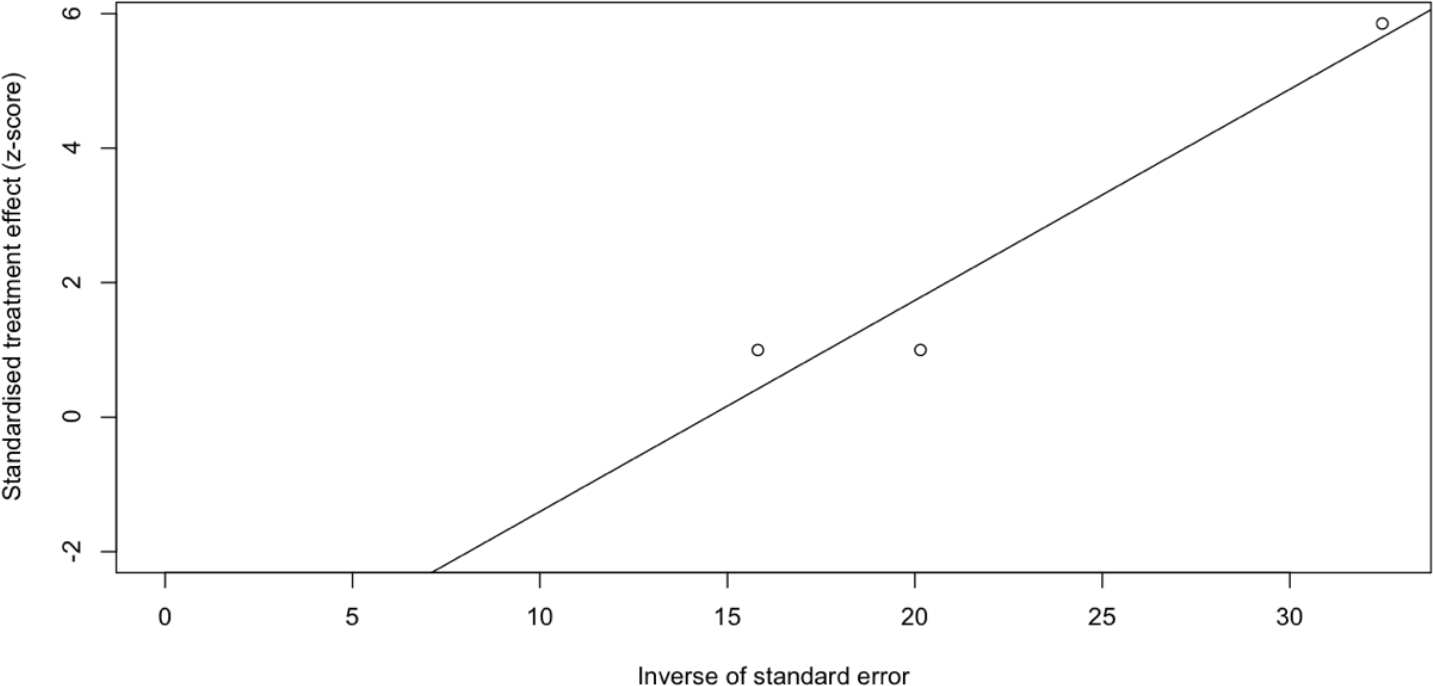
Egger’s publication bias plot (school staff infection attack rate)

### SARS-CoV-2 positivity rate

We also meta-analyzed SARS-CoV-2 positivity rates in schools (Table 4) from a total of six cross-sectional studies which included 639 SARS-CoV-2 positive cases in 6,682 participants tested [18-23]. The result of the random effect meta-analysis showed that the pooled SARS-CoV-2 positivity rate of total study participants was 8.00% (95% CI: 2.17%–16.95%) with substantial heterogeneity (I^2^ = 99.2%) (Figure 5A), but no evidence of publication bias (Figure 5B, 5C).

**Table 4:** SARS-CoV-2 positivity rate meta-analyses results.

|  | Number of studies | n (positive cases) | N (participants tested) | Positivity rate (%) | 95% CI | Cochrane Q | I<sup>2</sup> | Tau-square | P-Egger |
| --- | --- | --- | --- | --- | --- | --- | --- | --- | --- |
| Total | 6 | 639 | 6682 | 8.00 | 2.17-16.95 | 602.62 | 99.20 | 0.0285 | 0.6216 |
| Student | 6 | 401 | 4538 | 8.74 | 2.34-18.53 | 436.33 | 98.90 | 0.0315 | 0.4977 |
| School staff | 5 | 108 | 1043 | 13.68 | 1.68-33.89 | 211.72 | 98.10 | 0.0728 | 0.1286 |

CI: confidence interval

**Figure 5A:**
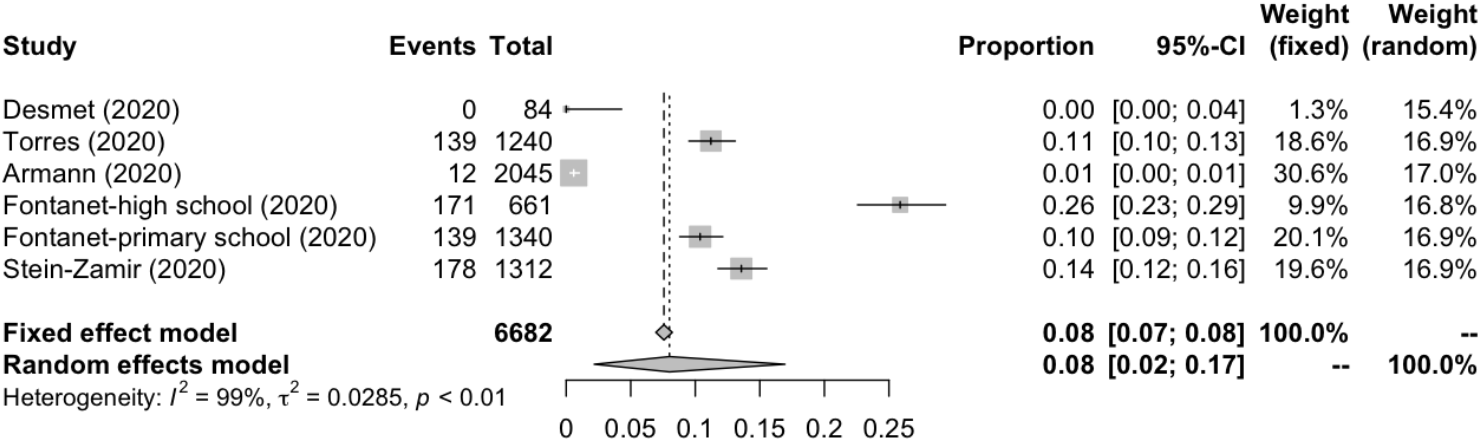
Forest plot (overall SARS-CoV-2 positivity rate)

**Figure 5B:**
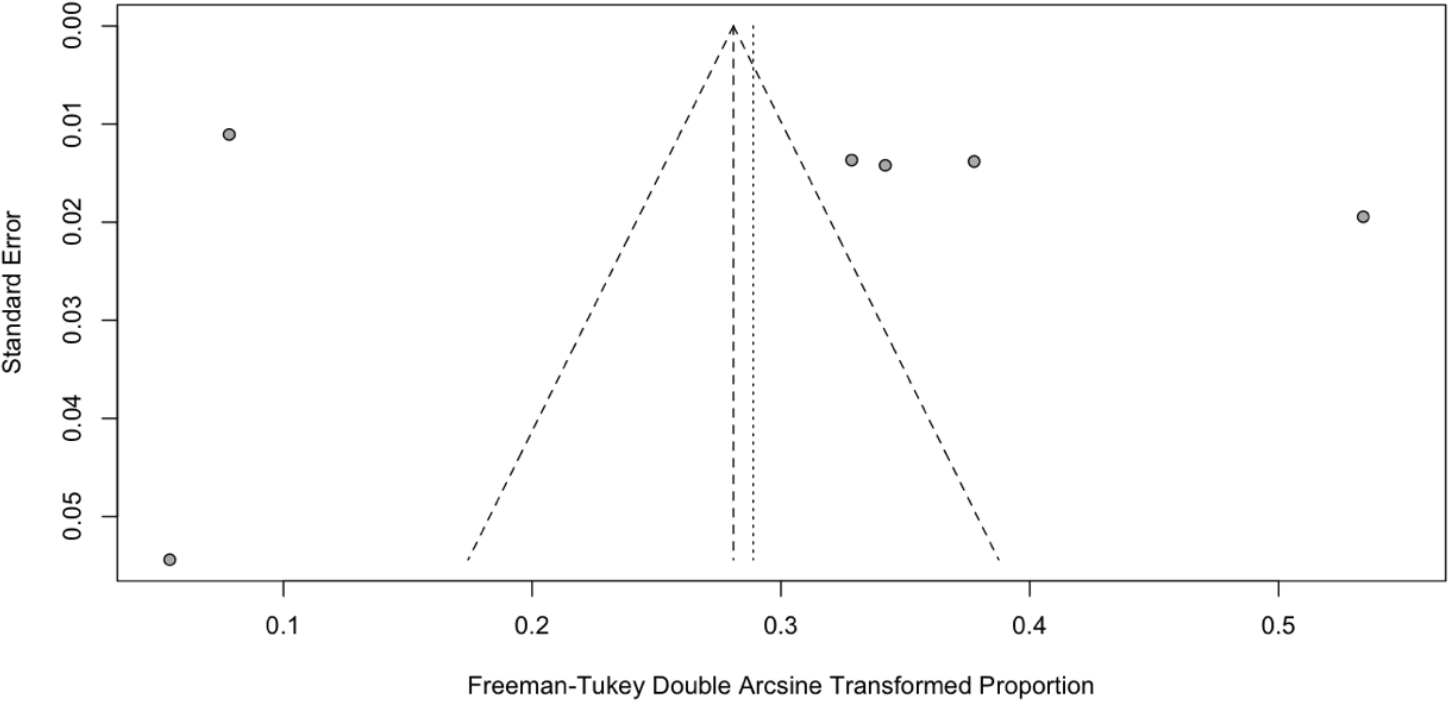
Funnel plot (overall SARS-CoV-2 positivity rate)

**Figure 5C:**
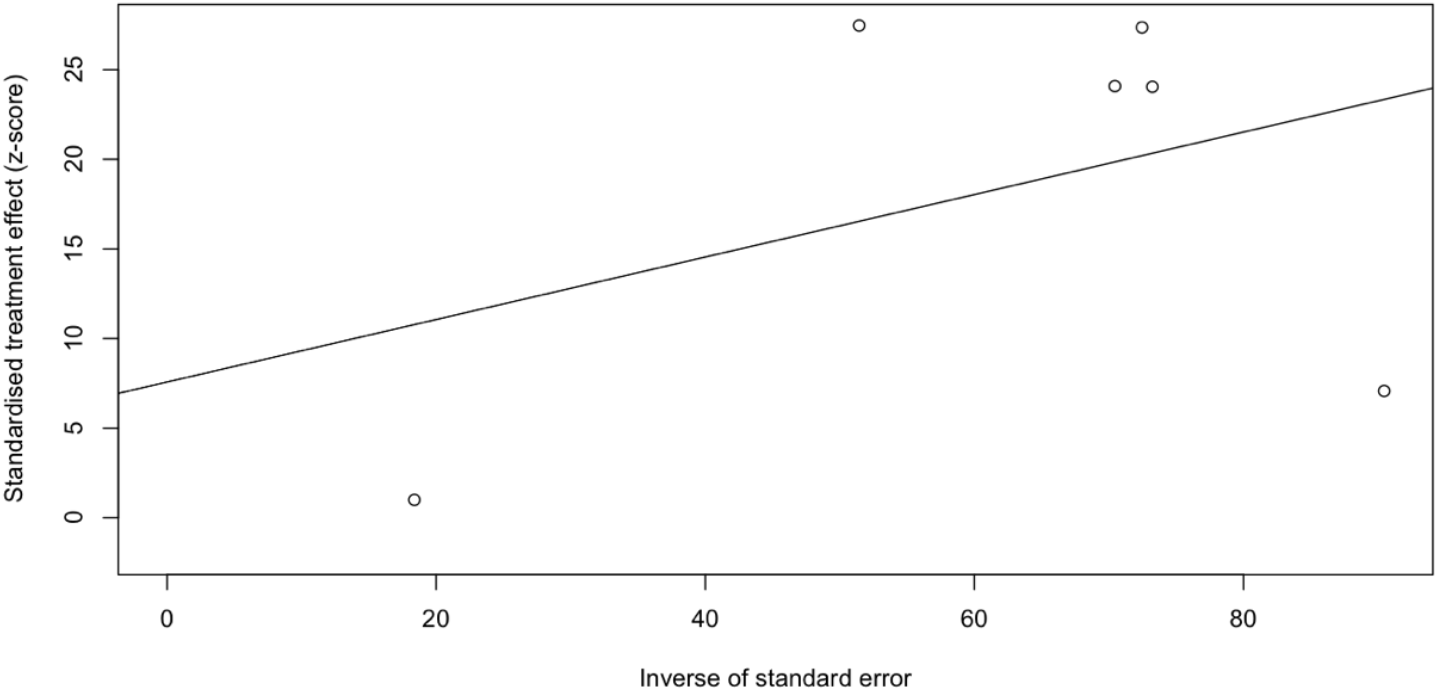
Egger’s publication bias plot (overall SARS-CoV-2 positivity rate)

Specifically, the positivity rates of SARS-CoV-2 was estimated to be 8.74% (95% CI: 2.34%–18.53%) among students (Figure 6A), compared to 13.68% (95% CI: 1.68%–33.89%) among school staff (Figure 7A). Heterogeneity was reported with I^2^ value of 98.9% and 98.1%. Funnel plot with Egger’s test (P = 0.498 and 0.129) suggested that there was no notable evidence of publication bias (Figure 6B, 6C and 7B, 7C).

**Figure 6A:**
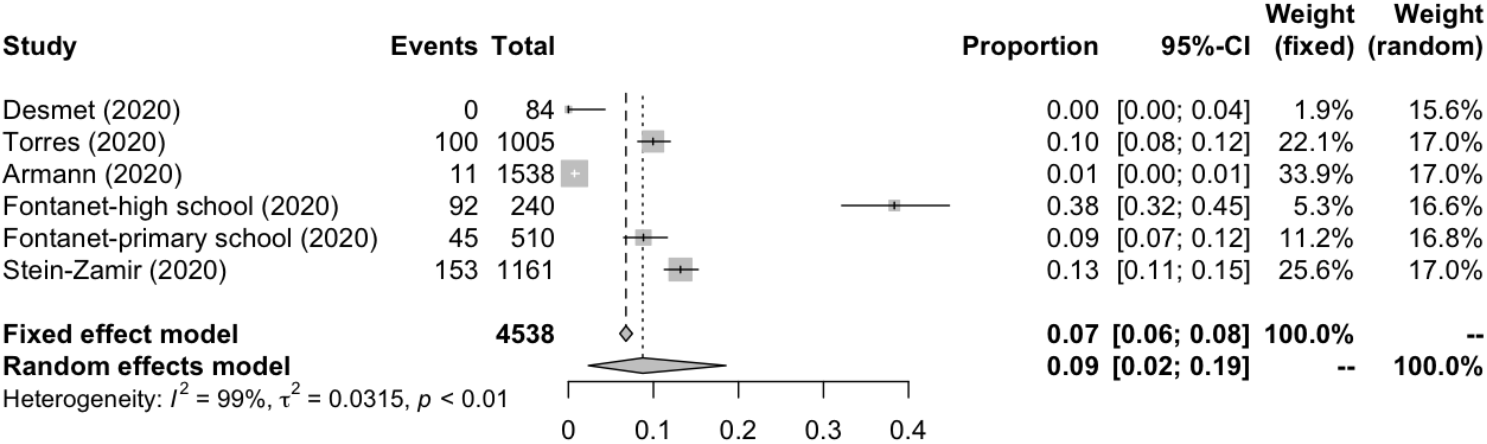
Forest plot (student SARS-CoV-2 positivity rate)

**Figure 6B:**
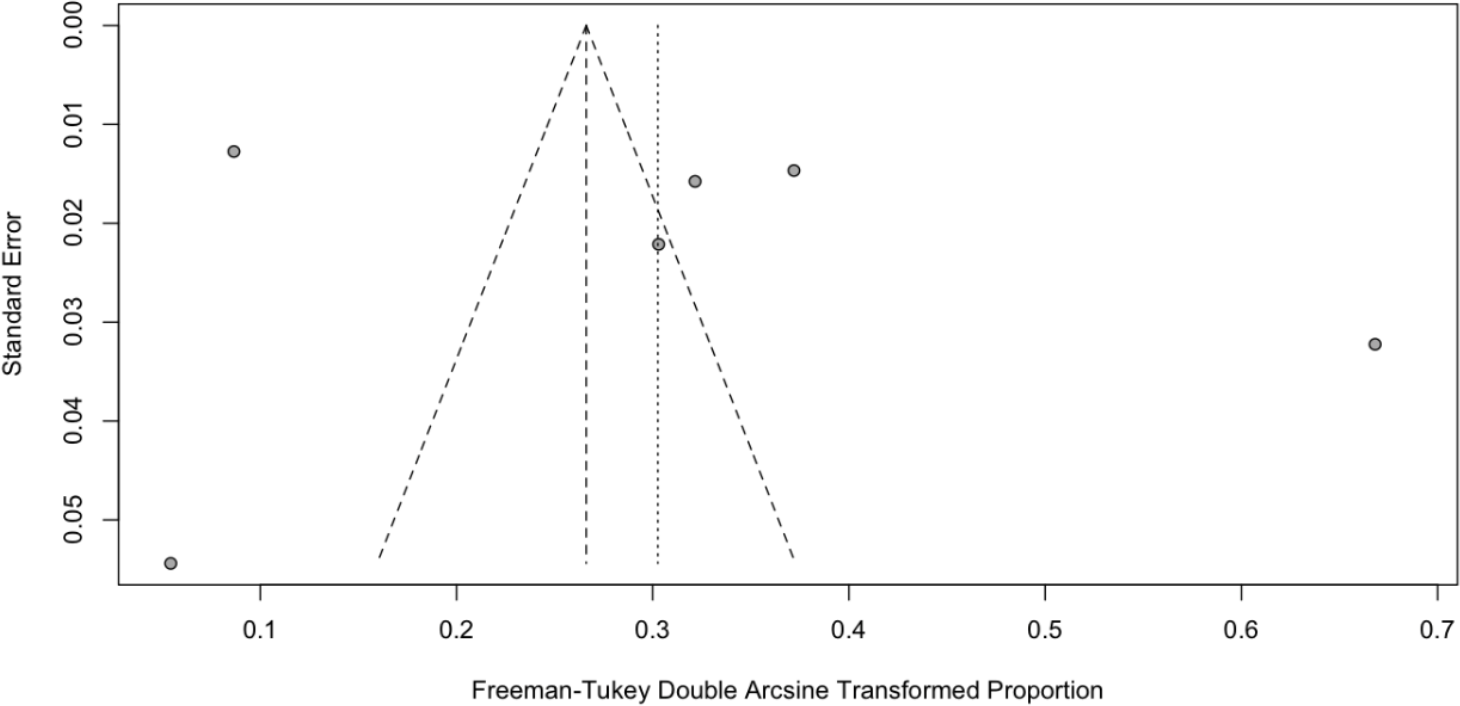
Funnel plot (student SARS-CoV-2 positivity rate)

**Figure 6C:**
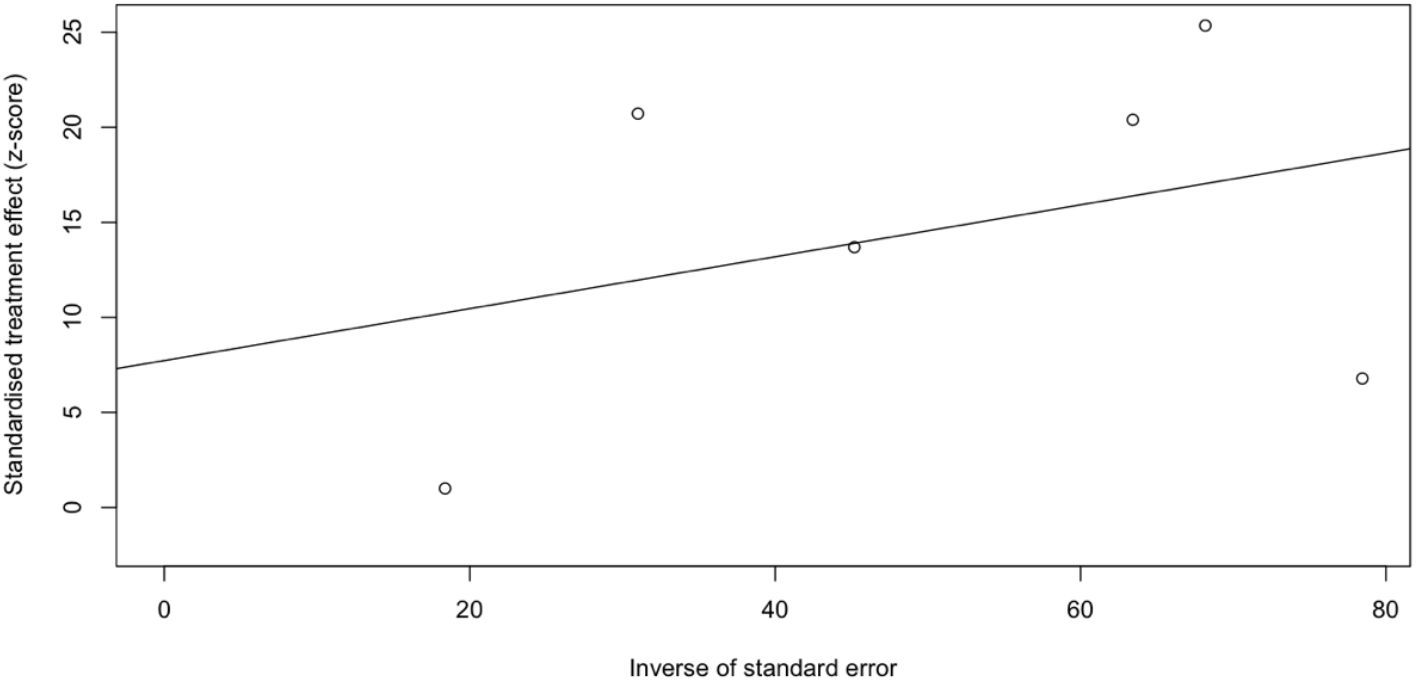
Egger’s publication bias plot (student SARS-CoV-2 positivity rate)

**Figure 7A:**
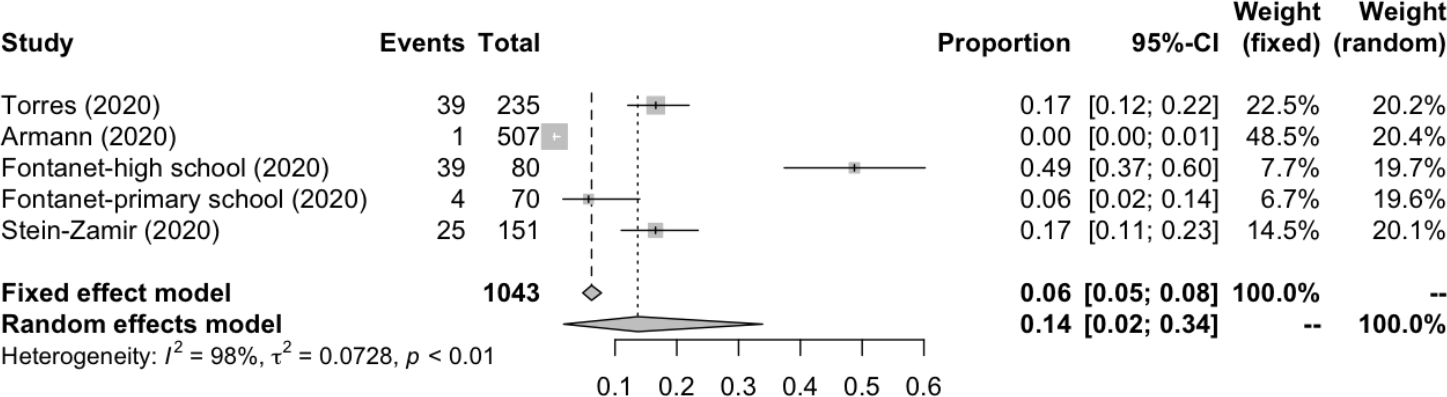
Forest plot (school staff SARS-CoV-2 positivity rate)

**Figure 7B:**
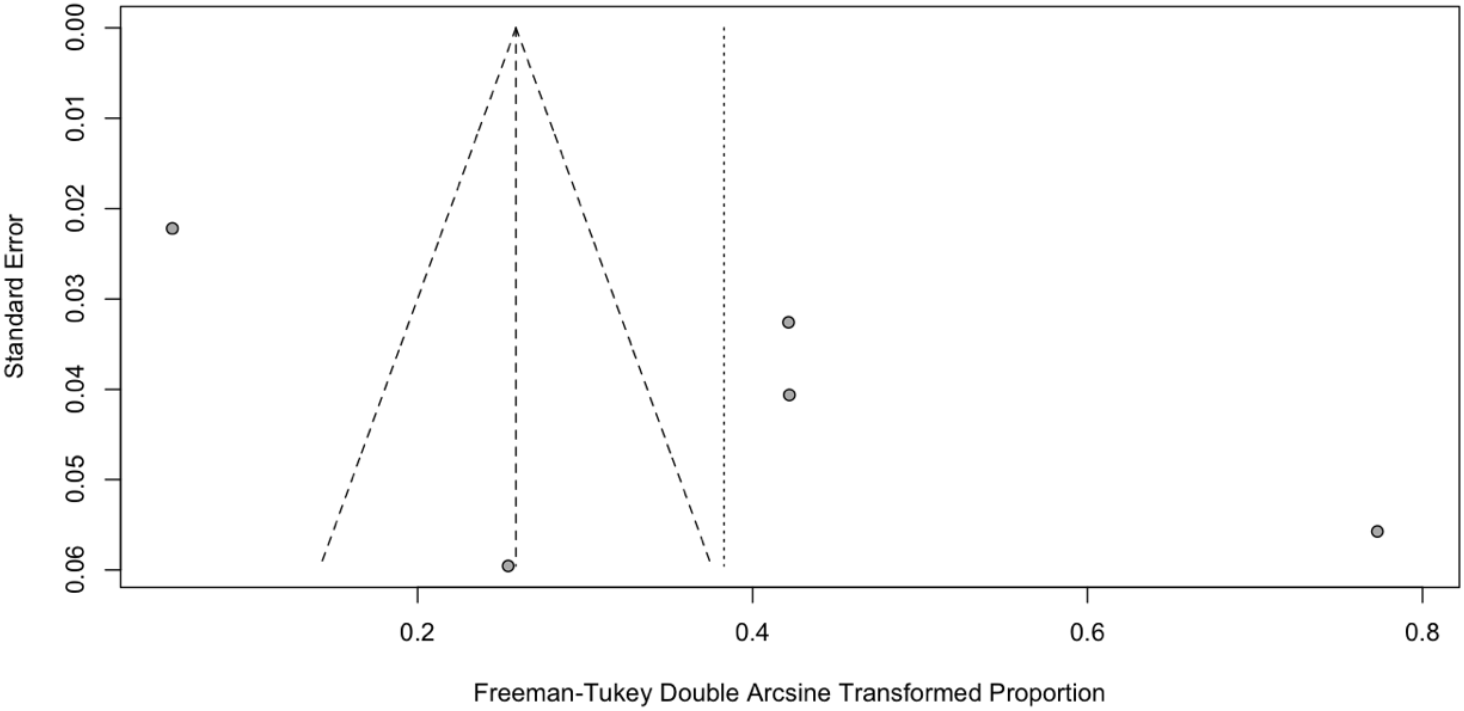
Funnel plot (school staff SARS-CoV-2 positivity rate)

**Figure 7C:**
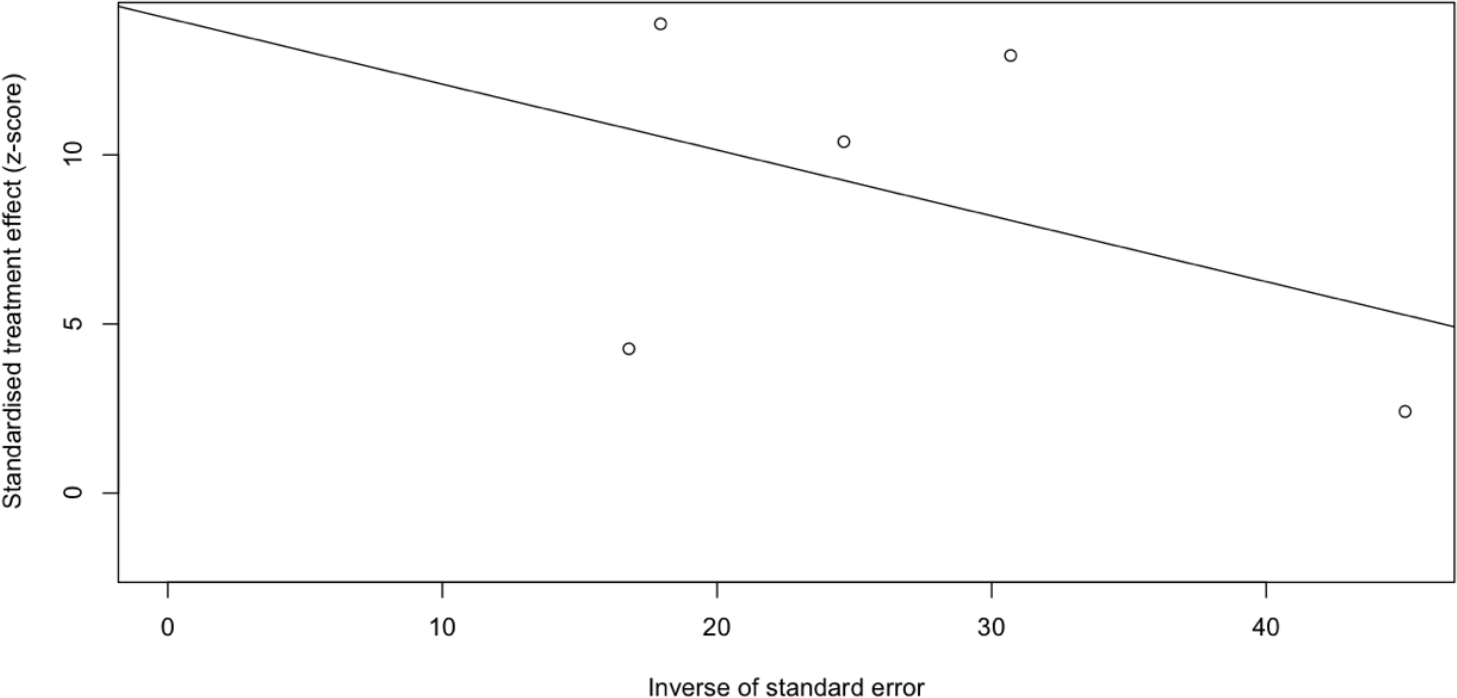
Egger’s publication bias plot (school staff SARS-CoV-2 positivity rate)

### Gender differences in SARS-CoV-2 infection attack rate and positivity rate

We did not find any gender differences for secondary infection (OR: 1.44, 95% CI: 0.50-4.14, P= 0.49, No. of cases in male/female group: 7/7) [17], and for SARS-CoV-2 positivity (OR: 0.90, 95% CI: 0.72-1.13, P= 0.36, No. of cases in male/female group: 268/359) in schools (Table 5) [18-19, 22-23].

**Table 5:**
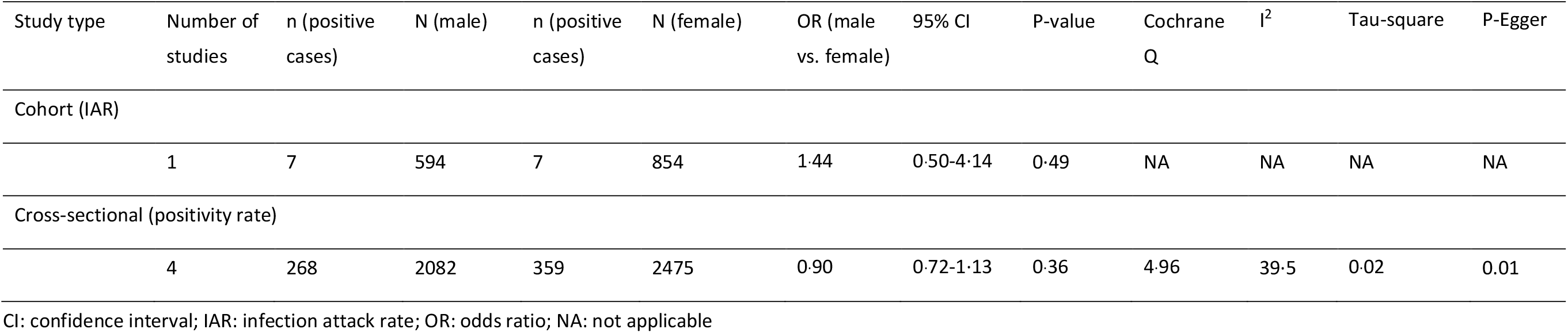
Gender differences in SARS-CoV-2 infection attack rate and positivity rate.

| Study type | Number of studies | n (positive cases) | N (male) | n (positive cases) | N (female) | OR (male vs. female) | 95% CI | P-value | Cochrane Q | I<sup>2</sup> | Tau-square | P-Egger |
| --- | --- | --- | --- | --- | --- | --- | --- | --- | --- | --- | --- | --- |
| Cohort (IAR) |  |  |  |  |  |  |  |  |  |  |  |  |
|  | 1 | 7 | 594 | 7 | 854 | 1.44 | 0.50-4.14 | 0.49 | NA | NA | NA | NA |
| Cross-sectional (positivity rate) |  |  |  |  |  |  |  |  |  |  |  |  |
|  | 4 | 268 | 2082 | 359 | 2475 | 0.90 | 0.72-1.13 | 0.36 | 4.96 | 39.5 | 0.02 | 0.01 |

CI: confidence interval; IAR: infection attack rate; OR: odds ratio; NA: not applicable

### Clinical symptoms

We also explored symptoms association with SARS-CoV-2 positivity (Table 6). Symptoms data was available from two sero-prevalence studies for both students and staff [22-23]. Study participants who had experienced major symptoms were more likely to test positive, compared to those who had had minor or no symptoms (27.09%, 10.98%, and 8.98%, respectively, P <0.001). Fever, cough, dyspnea, ageusia, anosmia, rhinitis, sore throat, headache, myalgia, asthenia, and diarrhoea were all associated with the detection of SARS-CoV-2 antibodies (Table 6). The most frequently reported symptoms were anosmia 84.27% (95% CI: 76.64-90.59%), ageusia 79.58% (95% CI: 58.86-94.50%), myalgia 30.61% (95% CI: 11.05-54.74%), fever 29.88% (95% CI: 8.32-57.73%), and diarrhoea 29.15% (95% CI: 8.74-55.32%).

**Table 6:** Clinical symptoms.

| Cross-sectional studies (N=2) |  |  |  |  |  |  |  |  |
| --- | --- | --- | --- | --- | --- | --- | --- | --- |
| Symptoms | n (positive cases) | N | Seropositivity rate (%) | 95% CI | P-value | Cochrane Q | I<sup>2</sup> | Tau-square |

| Symptom severity |  |  |  |  |  |  |  |  |  |
| --- | --- | --- | --- | --- | --- | --- | --- | --- | --- |
|  | None | 61 | 811 | 8.98 | 2.38-19.09 |  | 13.97 | 92.80 | 0.01 |
|  | Minor | 37 | 357 | 10.98 | 3.75-21.22 |  | 6.57 | 85.20 | 0.01 |
|  | Major | 212 | 833 | 27.09 | 10.23-48.36 | <0.001 | 40.17 | 97.50 | 0.02 |
| Ageusia | Yes | 93 | 118 | 79.58 | 58.86-94.50 | <0.001 | 6.16 | 83.80 | 0.02 |
|  | No | 217 | 1883 |  |  |  |  |  |  |
| Anosmia | Yes | 91 | 108 | 84.27 | 76.64-90.59 | <0.001 | 0.03 | 0.00 | 0.00 |
|  | No | 219 | 1893 |  |  |  |  |  |  |
| Asthenia | Yes | 135 | 513 | 27.79 | 13.02-45.55 | <0.001 | 17.27 | 94.20 | 0.02 |
|  | No | 175 | 1488 |  |  |  |  |  |  |
| Cough | Yes | 127 | 581 | 22.91 | 8.15-42.29 | <0.001 | 25.24 | 96.00 | 0.02 |
|  | No | 183 | 1420 |  |  |  |  |  |  |
| Diarrhoea | Yes | 65 | 238 | 29.15 | 8.74-55.32 | <0.001 | 16.70 | 94.00 | 0.03 |
|  | No | 245 | 1763 |  |  |  |  |  |  |
| Dyspnoea | Yes | 60 | 213 | 28.58 | 16.79-42.03 | <0.001 | 4.29 | 76.70 | 0.01 |
|  | No | 250 | 1788 |  |  |  |  |  |  |
| Fever | Yes | 127 | 461 | 29.88 | 8.32-57.73 | <0.001 | 36.22 | 97.20 | 0.04 |
|  | No | 183 | 1540 |  |  |  |  |  |  |
| Headache | Yes | 126 | 525 | 25.57 | 7.81-49.08 | <0.001 | 31.25 | 96.80 | 0.03 |
|  | No | 184 | 1476 |  |  |  |  |  |  |
| Myalgia | Yes | 109 | 366 | 30·61 | 11·05-54·74 | <0·001 | 22·59 | 95·60 | 0·03 |
|  | No | 201 | 1635 |  |  |  |  |  |  |
| Rhinitis | Yes | 117 | 506 | 22·37 | 6·72-43·70 | <0·001 | 27·56 | 96·40 | 0·03 |
|  | No | 193 | 1495 |  |  |  |  |  |  |
| Sore throat | Yes | 86 | 439 | 20·44 | 7·60-37·41 | 0·007 | 15·57 | 93·60 | 0·02 |
|  | No | 224 | 1562 |  |  |  |  |  |  |

CI: confidence interval

### Study quality

We considered contact-tracing studies as potential controlled cohort studies with the contacts of the index case representing the exposed group and the non-contacts who were in the school environment representing the unexposed group (a proxy community control group). Studies performed well in terms of representativeness of the groups and comparability. All studies employed active symptom screening in the exposed group with four of five studies employing passive or no screening in the unexposed groups and no testing. This difference in screening and testing introduces a risk of measurement bias. In a single study of three schools in Singapore [15], both the exposed and unexposed groups underwent PCR testing regardless of symptoms. Follow-up rates were reported for the exposed groups with less than 80% follow-up for all studies introducing a high risk of attrition bias across studies (Table 7).

**Table 7:** Quality assessment using the Newcastle-Ottawa Scale for cohort studies.

| Study ID | Selection Bias |  |  |  | Comparability |  | Detection Bias |  | Attrition Bias |  |
| --- | --- | --- | --- | --- | --- | --- | --- | --- | --- | --- |
|  | Representativeness of exposed group | Representativeness of unexposed group | Ascertainment of exposure | Outcome not present at start of study | Matching for school mitigation policies | Matching for age | Assessment of SARS-CoV-2 | Confirmation of SARS-CoV-2 | Adequacy of length of follow-up | Loss-to-follow-up rate |
| Macartney-2020 | * | * | ** | - | * | * | - | - | * | - |
| Danis-2020 | * | * | * | - | * | * | - | - | * | - |
| Heavey-2020 | * | * | - | - | * | * | - | - | * | - |
| Yung-2020 | * | * | * | - | * | * | - | * | * | - |
| NCIRS-2020 | * | * | * | - | * | * | - | - | * | - |

An '\*' denotes that the study met the criteria; a '-' denotes either that the study did not meet criteria or that it was not reported

For cross-sectional studies, we noted that while the sample for the target school population was representative, four out of six studies experienced poor response rates, introducing selection bias. Studies performed variably across other domains (Table 8).

**Table 8:** Quality assessment using a modified Newcastle-Ottawa Scale for cross-sectional studies.

| Study ID | Selection Bias |  | Performance Bias |  |  | Detection Bias |  |  | Attrition Bias | Comparability |  |
| --- | --- | --- | --- | --- | --- | --- | --- | --- | --- | --- | --- |
|  | Representativeness of sample | Percentage participation | Ascertainment of COVID_19 | Confirmation of COVID-19 | Blinding of assessors to prior exposure | Ascertainment of exposure to COVID-19 | Confirmation of exposure to COVID-19 | Blinding of assessors to COVID-19 status | Percentage in final analysis | Comparable in school | Comparable in age |
| Desmet-2020 | * | - | * | - | - | - | - | - | * | * | * |
| Armann-2020 | * | - | * | * | * | * | - | * | - | * | * |
| Fontanet-2020<br>Primary | * | - | * | - | * | * | - | * | * | - | - |
| Fontanet-2020<br>High | * | - | * | * | * | * | - | * | * | - | - |
| Torres-2020 | * | * | - | - | - | * | - | - | - | * | * |
| Stein-Zamir-2020 | * | * | * | - | - | * | - | - | * | * | * |

An '\*' denotes that the study met the criteria; a '-' denotes either that the study did not meet criteria or that it was not reported

## DISCUSSION

This systematic review summarizes the most recently available evidence to understand SARS-CoV-2 transmission in schools and includes an assessment of study quality to aid interpretation. The results from cohort and cross-sectional studies found that the overall IAR and SARS-CoV-2 positivity rate in school settings are low, and confirmed that students reported both lower IAR and SARS-CoV-2 positivity rate compared to school staff. However, the quality of studies limits our confidence in the observed results.

Cohort studies reported limited evidence of SARS-CoV-2 transmission in school settings. Compiling the data from five studies of school exposures early in pandemic before lockdown, we report an overall IAR of 0.08% (95% CI: 0.00%–0.86%). Clusters in educational facilities were identified in one of the five reporting countries, and those that occurred were limited in number and size [17]. NSW did not close schools during the beginning of COVID-19 pandemic. Transmission rates of student-to-student, student-to-staff, staff-to-student and staff-to-staff were 0.31%, 0.97%, 1.49% and 4.38% respectively. Students reported lower IAR than school staff. In addition, there is uncertainty about in which grade school children are more likely susceptible to and transmit SARS-CoV-2. IARs for ECDC (<6 years old), primary school (6-12 years old), and secondary school (12-18 years old) were 2.25%, 0.92%, 0.00% respectively in NSW. The data is limited to reach a consensus. However, the clusters in NSW demonstrated that classroom crowding and other factors related to physical distancing may play a role in the spread of SARS-CoV-2 under the school environment.

Many countries such as Denmark, Austria, Finland, Norway have implemented various prevention and control measures [25] and those countries have smaller classroom sizes compared to Australia [26]. The remaining four studies in France (Les Contamines-Montjoie), Ireland, Singapore, Australia (NSW) reported that transmission rate from student-to-student, student-to-staff, staff-to-student and staff-to-staff was 0.00% [13-16]. The limited evidence available to quantify the extent of SARS-CoV-2 transmission in school settings, reflects the fact that cluster outbreaks have been studied and reported relatively infrequently in schools to date. Effective implementation of NPIs such as physical distancing, small-size class, cancellation of mass gatherings, and hand hygiene is likely to further limit our ability to study school transmission [6].

Cross-sectional studies estimated the proportion of SARS-CoV-2 positive cases, to give an insight into how many people have been infected in schools. As described above, the positivity results in the general study population under school environment varied from 0.00 (lowest in eight daycare centers in Belgium) to 25.87% (highest in one high school in France), which is likely to reflect the community positivity rate at the time the study was conducted [18-23]. The lower positivity rate in students suggested that students are less susceptible to infection and/or less frequently infected than adult school staff, which indicated that students are not at higher risk of causing super-spreading events in schools. Our finding is in line with many previous studies comparing sero-prevalence between children and adults [27-31]. However, the quality of the included studies is low and we should interpret the result with caution. Sero-prevalence results from Sweden in which schools remained open, demonstrated that 5-19 year olds (6.0%, 95% CI 2.3-10.2%) children had similar sero-prevalence to 20-49 year olds (8.5%, 95%CI 4.99-11.7) adults [32]. We suggest more specialised and large-scale sero-surveillence studies need to be conducted to monitor SARS-CoV-2 infection during school opening. In addition, there is no consensus about in which grade school children have higher susceptibility to SARS-CoV-2 Infection. SARS-CoV-2 positive rates for pre-school (<6 years old), primary school (6-12 years old), middle school and high school (12-18 years old) were 12.24%, 10.84%, 11.80% and 5.69% in Chile (Santiago). The peak rate was observed in pre-school. By comparison, SARS-CoV-2 positive rates were 8.82% in primary schools (6-12 years old) in France (Crépy-en-Valois), and were 0.72% in middle schools (12-16 years old) in Germany (Saxony), and 38.33% in high schools (12-18 years old) in France, 13.18% in high schools (12-18 years old) in Israel.

A single study in Israel investigated whether school transmission increased relative to community transmission. Compared with the school-closure period, the total number of COVID-19 cases increased, and the proportion of infected children increased from 19.8% to 40.9% in the community. However, the role of school in the significant COVID-19 increase in the community was unclear because school re-opening coincided with the relaxation of other prevention and control measures [18].

We did not find any gender differences for secondary infection and SARS-CoV-2 positivity in schools. The lack of sex-disaggregated data for student and school staff specifically in the reviewed studies enhanced the difficulty to further explore potential explanations for the findings in gender.

The main clinical symptoms for COVID-19 patients were anosmia (84.3%), ageusia (79.9%), myalgia (30.6%), fever (29.9%), diarrhoea (29.2%), dyspnea (28.6%), and cough (22.9%). We should interpret the result with caution because the symptom data only come from two sero-surveillance studies carried out in one high school (n=661) and six primary schools (n=1340) in France (Crépy-en-Valois). Studies from Italy [33-35], Germany [36], UK [37], Turkey [38] and Sweden [39] also reported similar clinical symptoms in children. In addition to common respiratory symptoms, gastrointestinal symptoms such as diarrhoea were present in around 25% of pediatric patients [24]. It is noted that the persistent shedding of SARS-CoV-2 in stools of infected children has been consistently reported, showing that SARS-CoV-2 may be present in the gastrointestinal tract for a longer duration than viral presence in the respiratory system, compared to adults [40-42].

The main strength of this study is that it provides a critical assessment of the published epidemiological evidence on SARS-CoV-2 transmission risk in the school environment. In addition, we estimated pooled IARs and SARS-CoV-2 positivity rates for students and school staff, and to our best knowledge, this is the first meta-analyses conducted, to investigate what is the rate of SARS-CoV-2 transmission in schools. However, the following potential limitations should be considered. First, further interpretation of age-group differences in IARs and positivity rates could not be performed because 80.0% (4/5) of included cohort studies and 50.0% (3/6) of included cross-sectional studies did not specify the ages of students and school staff. The remaining four included studies did not provide the raw data and we could not unify different age groups to run the meta-analysis. Second, cross-comparisons between IARs and positivity rates reported in different regions/countries is difficult because of differences in the sampling and testing methods used, timing of the studies in relation to the outbreak, response measures and underlying community transmission. Moreover, the differences may contribute to the heterogeneity observed in the meta-analyses results and raise methodological concerns around the validity of the meta-analysis. Due to the limited number of included studies, we could not conduct subgroup meta-analyses to further investigate the heterogeneity. As this is a living review, we anticipate that with the addition of more, well-conducted studies over time, heterogeneity may improve. Third, only two studies in the included 11 studies (18.2%) reported prevention and control measures in place in schools such as class size, physical distancing, and staggered class start and end times, making it difficult to further investigate the effectiveness of NPIs under the school environment. Forth, only one study (9.1%) compared school transmission rate with community transmission rate. Few studies have assessed the impact of school opening on transmission outside the school. Thus, we additionally searched study’s background sero-prevalence or SARS-CoV-2 case rate per 100,000 population online, however, the data is limited. We suggest future studies could investigate this research question: does school increase or decrease transmission to the community. Fifth, although there is no evidence for publication bias, the number of included studies were less than ten. When there are fewer studies, the power of the tests is too low to distinguish chance from real asymmetry. Therefore, results should be interpreted with caution. Lastly, the majority of included studies are pre-print publications and have not been peer-reviewed. The quality of the included studies is low and we should interpret the results with caution.

## CONCLUSION

In conclusion, although there is limited evidence available to quantify the extent of SARS-CoV-2 transmission in schools, the balance of evidence so far indicates that the overall IAR and SAR-CoV-2 positivity rate in the school environment are low. Specifically, lower IAR and positivity rates were reported in students compared to school staff, but poor study quality limits our overall confidence in these results. However, it is important to implement effective NPIs such as physical distancing, small-size class to prevent schools from becoming a setting for accelerating onward transmission during the re-opening of schools.

## Data Availability

All data generated or used during the study appear in the submitted article.

## Acknowledgements

UNCOVER (Usher Network for COVID-19 Evidence Reviews) authors that contributed to this review are: Prof Harry Campbell, Dr Ruth McQuillan, Prof Harish Nair, Ms Emilie McSwiggan, Prof Gerry Fowkes. https://www.ed.ac.uk/usher/uncover.

## Authorship contributions

The UNCOVER group conceived this study. XL, WX, MD, YH, ZL and AK conducted literature review. WX and XL performed meta-analyses. NS and CM developed the quality assessment tools and conducted quality assessment. WX and XL wrote the draft of the paper with input from all co-authors. ET, NS and CM provided methodological guidance on conducting the review. All authors have read and approved the final manuscript as submitted.

