## Supplementary Materials for "What is the evidence for transmission of COVID-19 by children in schools? A living systematic review"

### Appendix

#### Search Strategy:

**MEDLINE(R) (Ovid)**

Date of search: 2020-07-14 and 2020-09-14

1. schools/ or schools, nursery/
2. exp child day care centers/ or nurseries, infant/
3. school*.mp.
4. preschool*.mp.
5. (nurseries or nursery).mp.
6. kindergarten*.mp.
7. (day care or daycare).mp.
8. playgroup*.mp.
9. 1 or 2
10. 3 or 4 or 5 or 6 or 7 or 8
11. 9 or 10
12. exp Betacoronavirus/
13. exp Coronavirus Infections/
14. exp Coronavirus/
15. (2019nCoV or Betacoronavirus* or Corona Virus* or Coronavirus* or Coronovirus* or CoV or CoV2 or COVID or COVID19
16. or COVID-19 or HCoV-19 or nCoV or SARS CoV 2 or SARS2 or SARSCoV or SARS-CoV or SARS-CoV-2 or Severe Acute Respiratory Syndrome CoV*).mp. [mp=title, abstract, original title, name of substance word, subject heading word, floating sub-heading word, keyword heading word, organism supplementary concept word, protocol supplementary concept word, rare disease supplementary concept word, unique identifier, synonyms]
17. 12 or 13 or 14 or 15
18. 11 and 16
19. (201912* or 2020*).ed.
20. 17 and 18

**CINAHL (EBSCO)**

Date of search: 2020-07-14

| | |
|---|---|
| S8 | S7 AND EM202012- |
| S7 | S5 AND S6 |
| S6 | S3 OR S4 |
| S5 | S1 OR S2 |

S4 2019nCoV or Betacoronavirus* or Corona Virus* or Coronavirus* or Coronovirus* or CoV or CoV2 or COVID or COVID19 or COVID-19 or HCoV-19 or nCoV or SARS CoV 2 or SARS2 or SARSCoV or SARS-CoV or SARS-CoV-2

S3 (MH "Coronavirus Infections+") OR (MH "Coronavirus+")

S2 school* OR highschool* OR nursery OR nurseries OR preschool* OR Kindergarten* OR "day care" OR daycare OR playgroup

S1 (MH "Schools") OR (MH "Schools, Elementary") OR (MH "Schools, Middle") OR (MH "Schools, Nursery") OR (MH "Schools, Secondary") OR (MH "Schools, Special") OR (MH "Child Day Care")

**ERIC (ProQuest)**

Date of search: 2020-07-14

2019nCoV OR Betacoronavirus* OR "Corona Virus*" OR Coronavirus* OR Coronovirus* OR CoV OR CoV2 OR COVID OR COVID19 OR COVID-19 OR HCoV-19 OR nCoV OR "SARS CoV 2" OR SARS2 OR SARSCoV OR SARS-CoV OR SARS-CoV-2

**Embase (Ovid)**

Date of search: 2020-07-14

1. school/ or high school/ or kindergarten/ or middle school/ or nursery school/ or primary school/
2. day care/
3. (school* or highschool* or preschool* or nurseries or nursery or kindergarten* or day care or daycare or playgroup*).mp. [mp=title, abstract, heading word, drug trade name, original title, device manufacturer, drug manufacturer, device trade name, keyword, floating subheading word, candidate term word]
4. exp Coronavirinae/
5. exp Coronavirus infection/
6. (Corona virus or Corona Virus* or Coronavirus* or Coronovirus* or CoV or CoV2 or COVID or COVID19 or COVID-19 or HCoV-19 or nCoV or SARS CoV 2 or SARS2 or SARSCoV or SARS-CoV or SARS-CoV-2 or 2019nCoV).mp. [mp=title, abstract, heading word, drug trade name, original title, device manufacturer, drug manufacturer, device trade name, keyword, floating subheading word, candidate term word]
7. Betacoronavirus*.mp.

8. 1 or 2 or 3

9. 4 or 5 or 6 or 7

10. 8 and 9

11. ("201948" or "201949" "20195*" or 2020*).em.

12. 10 and 11

**WHO COVID-19 database** via https://search.bvsalud.org/global-literature-on-novel-coronavirus-2019-ncov/

Date of search: 2020-07-14

tw:(school* OR highschool* OR preschool* OR daycare OR "day care" OR nursery OR nurseries OR kindergarten* OR playgroup* OR playschool*)

**medRxiv** via https://mcguinlu.shinyapps.io/medrxivr/

Date of search: 2020-07-14

[Setting terms combined internally with OR and with the COVID group using AND]

[Ss]chool

[Hh]ighschool

[Pp]reschool

[Nn]urseries

[Nn]ursery

[Kk]indergarten

[Dd]ay care

[Dd]aycare

[Pp]laygroup

[COVID terms combined internally with OR and with the Setting group using AND]

2019nCoV

Betacoronavirus

Corona Virus

Coronavirus

Coronovirus

\\bCoV\\b

\\bCoV2\\b

COVID

HCoV-19

\\bnCoV\\b

SARS CoV 2

SARS2

SARSCoV

SARS-CoV

Grey literature searches

Date of search: 2020-07-14

These searches were conducted using Google Advanced search within the domains of selected organisations.

Required term: school

Any of these: 2019nCoV Betacoronavirus "Corona Virus" Coronavirus Coronovirus CoV CoV2 COVID COVID19 COVID-19 HCoV-19 nCoV "SARS CoV 2" SARS2 SARSCoV SARS-CoV SARS-CoV-2

Site domains searched:

- American Academy of Pediatrics: https://www.aap.org
- The Royal College of Paediatrics and Child Health (RCPCH): https://www.rcpch.ac.uk/
- Don’t forget the bubbles: https://dontforgetthebubbles.com/

**NEWCASTLE - OTTAWA QUALITY ASSESSMENT TOOL**

**COHORT STUDIES**

Note: A study can be awarded a maximum of one star for each numbered item within the Selection and Outcome categories. A maximum of two stars can be given for Comparability

### Selection

1) Representativeness of the exposed cohort

   a) truly representative of the average school community ✵
   b) somewhat representative of the average school community ✵
   c) selected group of school-going children and staff
   d) no description of the derivation of the cohort

2) Selection of the non-exposed cohort

   a) drawn from the same school or community as the exposed cohort ✵ (award ✵ if non-exposed group is defined by status of not being a close contact of the index case if not formally reported as non-exposed)
   b) drawn from a different source
   c) no description of the derivation of the non-exposed cohort

3) Ascertainment of exposure

   a) school review of time-tables ✵
   b) structured interview ✵
   c) self-report
   d) no description

4) Demonstration that outcome of interest (COVID-19) was not present at start of study

   a) yes ✵
   b) no

### Comparability

1) Comparability of cohorts on the basis of the design or analysis

   a) study controls for school non-pharmaceutical interventions ✵

b) study controls for age ✵

**Outcome**

1) Assessment of outcome

a) active symptom screening in exposed and non-exposed group ✵

b) active symptom screening in exposed and passive follow-up in non-exposed group

c) passive symptom screening in both exposed and non-exposed groups

d) no description

2) How was outcome (COVID-19) confirmed?

a) PCR in both groups ✵

b) Ab test in both groups if/when Ab test can detect current infection status ✵

c) independent duplicate assessment of symptoms in both groups ✵

d) PCR/Ab/Duplicate screening in exposed group only

e) no description

3) Was follow-up long enough for outcomes to occur

a) yes (for 14 days or more) ✵

b) no (for less than 14 days)

4) Adequacy of follow up of cohorts

a) complete follow up - all participants accounted for ✵

b) participants lost to follow up unlikely to introduce bias - > 80 % follow up, or description provided of those lost ✵

c) follow up rate < 80% and no description of those lost

d) not described

**QUALITY ASSESSMENT TOOL**

**CROSS-SECTIONAL STUDIES**

Note: A study can be awarded a maximum of one star for each numbered item.

### Selection

1) Representativeness of the sample for the target population (school)

a) truly representative of the average school community ✵

b) somewhat representative of the average school community ✵

c) selected group of school-going children and staff

d) no description of the derivation of the cohort

2) What percentage of selected individuals agreed to participate?

a) 80 – 100% ✵

b) < 80%

c) no description

### Performance or Measurement

1) How was COVID-19 ascertained?

a) Naso- or oral-pharyngeal PCR ✵

b) Sero-Ab test ✵

c) Self-test for Ab using rapid test

d) questionnaire

e) no description

2) How was COVID-19 confirmed?

a) sero Ab test ✵

b) not confirmed

c) no description

3) Were assessors of COVID-10 blinded to prior exposure to possible case?

a) blinded stated or laboratory assessment ✵

b) not blinded

c) no description

**Detection**

1) How was prior exposure to contact with a case with COVID-19 ascertained?

a) Record checking on contact database e.g. register of notifiable disease ✹

b) Interview with school and timetable checking ✹

c) questionnaire

d) no description

2) How was exposure to a case with COVID-19 confirmed?

a) laboratory lists or register (if different to methods in Detection 1) ✹

b) questionnaire ((if different to methods in Detection 1) ✹

c) no description

3) Were assessors of prior exposure status blinded to COVID-19 results?

a) blinded stated or laboratory assessment ✹

b) questionnaire pre-dated COVID-19 test results

c) not blinded

d) no description

**Attrition**

1) Percentage of participants included in final analysis

a) complete follow up - all participants accounted for ✹

b) participants lost to follow up unlikely to introduce bias - > 80 % follow up, or description provided of those lost ✹

c) follow up rate < 80% and no description of those lost

d) not described

**Comparability (2 stars can be given)**

1) Comparability of cohorts on the basis of the design or analysis

a) study controls for school non-pharmaceutical interventions □

b) study controls for age
